# Temporal relationships between distress and pain in people living with HIV

**DOI:** 10.64898/2026.07.15.26358133

**Authors:** Gwendoline Arendse, Peter Kamerman, Antonia Wadley, Robert R Edwards, John Joska, Romy Parker, Victoria J Madden

## Abstract

**Objective:** There is a bidirectional relationship between emotional distress and pain. However, this relationship is understudied in people with HIV in low-resource settings. This study sought to describe the temporal relationship between emotional distress and pain in people with HIV.

**Design:** Longitudinal observational study.

**Methods:** Participants with virally suppressed HIV, who had previously reported either no pain or persistent pain, provided weekly remote ratings of distress, worst pain, and average pain using 0-10 visual analogue scales. Within-individual fluctuations in distress and pain were visualised over time. Cumulative link mixed models assessed concurrent and one-week-lagged associations between distress and worst or average pain.

**Results:** 72 participants provided responses over 49 weeks. The participants had a median (IQR) age of 43 (37–51) years, 63% (n=45) were unemployed and most were females (n=51;71%). Distress was associated with concurrently reported worst pain and average pain intensity (odds ratios, ORs=1.85-2.08; based on 195 response pairs from 30 participants). Time-lagged analyses (146 response pairs from 27 participants) showed worst pain (OR=1.42, 95% CI=1.17-1.71, p<0.001) and average pain (OR=1.43, 95% CI=1.20-1.71, p<0.001) intensity to be associated with higher distress in the next week. Distress was associated with higher worst pain intensity (OR=1.25, 95% CI=1.07-1.46, p=0.023) but not average pain intensity in the next week.

**Conclusions:** The temporal association between distress and worst pain intensity was bidirectional, whereas distress was not associated with average pain intensity in the next week. Both pain and emotional distress should receive attention from HIV research and clinical care in similar settings.

## Introduction

Approximately 40% of people with chronic pain have symptoms of depression and/or anxiety (1), and approximately 33% of people with common mental disorders (depression and anxiety) report pain (2). However, exactly how pain and emotional distress influence each other over time remains unclear.

Much research has studied *pain-focused* distress, investigating how various forms of psychological, emotional, and/or cognitive distress influence pain – especially, pain-focused worrying (e.g., ‘pain catastrophising’ (3), comprising constructs of magnification, rumination, and helplessness) and fear of pain, both of which are included in the cyclical fear-avoidance model of chronic pain (4, 5). Many cross-sectional and prospective studies link greater ‘pain catastrophizing’ to higher pain intensity, although some cross-sectional and prospective studies have not replicated this association (6–11).

Pain intensity may also weakly predict pain catastrophizing cross-sectionally (12, 13), but longitudinal findings are conflicting (14–17). Fear of pain is positively associated with clinical pain intensity in cross-sectional (7, 18) and longitudinal studies of pain after surgery and at work (19, 20). Fear-related avoidance of pain-eliciting activities also cross-sectionally predicts pain intensity (21–24), but longitudinal findings conflict again, across timescales up to 3 years (20).

Less research has probed how distress that is *not focused on pain* (here we use the umbrella term ‘general distress’), relates to pain, although plausible mechanisms exist, including prolonged dysregulation of the hypothalamic pituitary axis or threat-biased learning processes (for review, see 25). Cross-sectional studies have often found minimal relationship between general distress and pain (13, 24, 26, 27), whereas longitudinal studies suggest that distress prospectively predicts pain some months later – possibly more consistently than pain predicts later distress (6, 28). In the surgical field, higher patient-reported ‘general distress’ before surgery predicted more pain after surgery, both in the short term (29, 30) and some weeks and months later (31, 32), although there are exceptions (33–35). Crucially, however, most non-surgical longitudinal studies have used reasonably long time frames, which cannot resolve whether distress and pain influence each other over shorter timescales. One study using daily assessments in a small sample (n=8) found that the direction of the temporal relationship between pain and distress varied between individuals (36).

There is a particular need to probe the temporal relationships between non-pain-focused distress and pain in people with HIV, especially in the sub-Saharan African context that homes most of this population. People with HIV carry a high burden of both pain and general distress, alongside socio-economic factors that are themselves associated with both distress and pain (16, 37–40). This burden is underscored by one direct comparison between people with HIV and people with cancer, which found more pain interference and a higher burden of anxiety and depression in those with HIV (41). Pain catastrophising was also higher in the group with HIV, aligning with other reports of pain catastrophizing, fear of pain exacerbation, and indications of fear avoidance in people with HIV and chronic pain (41–45). Overlaps between general negative affective states, pain, and pain-focused anxiety are also relevant to pain in HIV, given that pain-focused anxiety is associated with symptoms of anxiety and depression (46), and general anxiety and general distress is associated with more severe pain, in people with HIV (16, 41, 43, 47), albeit inconsistently (48).

The current work was motivated by the need to better understand shared and overlapping mechanisms between general distress and pain, as a first step towards designing pragmatic, resource-efficient interventions for future clinical testing in sub-Saharan African people with HIV. We aimed to track week-to-week fluctuations in emotional distress and pain and to assess temporal associations between distress and pain intensity, in people with HIV who had previously reported chronic or no pain. We tested three possibilities: that distress and pain co-fluctuated simultaneously, that changes in distress preceded changes in pain, and that changes in pain preceded changes in distress (by one week).

## Methods

### Study overview

This prospective observational study in South Africa was attached to a larger parent study, which aimed to recruit 100 adults with HIV who had viral suppression and self-reported no pain or persistent pain (pragmatic sample size) (49), and assessed distress, pain, psychosocial and medical constructs. For the current dataset, participants provided additional written informed consent to remotely answer questions on distress and pain each week for 6 months, starting at varying time points relative to the parent study period. Study staff and participants were unacquainted with the research hypotheses; blinding was not assessed.

### Participants

Parent study participants were consenting adults aged 18-65 with viral load<50 copies/ml, no major psychiatric symptoms, and self-reporting no pain or persistent pain. The University of Cape Town Faculty of Health Sciences Human Research Ethics Committee (approval number: 764/2019) and the City of Cape Town (ref: 24699) provided ethical approval.

### Procedures

Data were collected from 16 March 2021 to 31 March 2022. Participants used a mobile phone app (Upinion [Opinion Solutions (Pty) Ltd]) that prompted users to answer 1-2 weekly questionnaires in their preferred language (English or isiXhosa). One questionnaire addressed pain and distress. From 30 August 2021, funding support allowed dedicated recruitment staff, study phones, and small incentives to support response rates (details in supplementary file).

### Study variables

Figure 1 summarises the questions posed to participants; Figure S1 shows the full version.

**Fig. 1.**
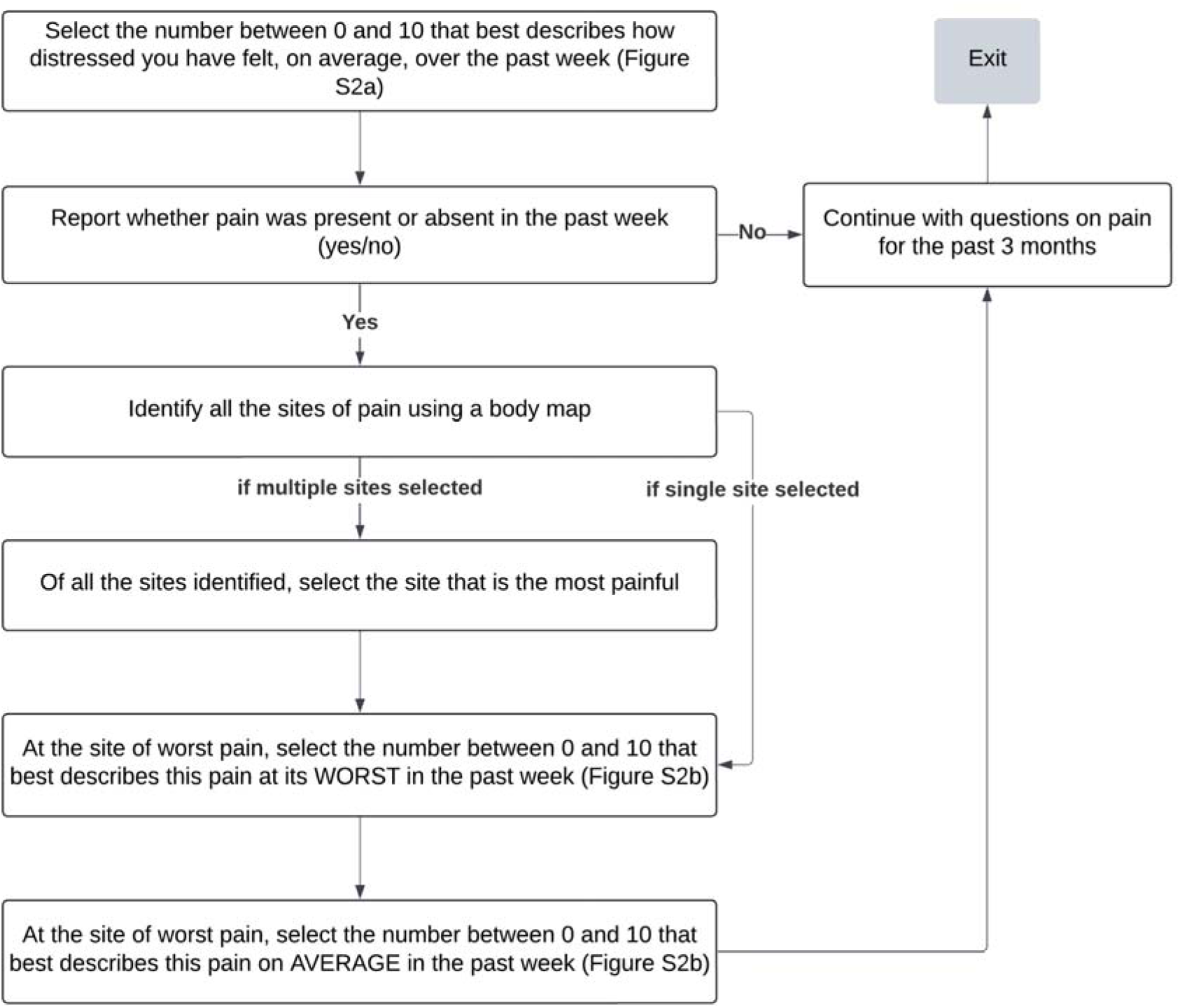
Overview of questions posed to participants; full details in Figure S1.

#### Distress

After choosing a preferred language and entering their participant study identifier, participants viewed a vertical numerical rating scale (NRS) alongside the instruction, “Throughout our lives, most of us feel distressed from time to time. Select the number between 0 and 10 that best describes how distressed you have felt, on average, over the past week.” NRS anchors were 0 (not distressed at all; bottom end) and 10 (as distressed as I could possibly be; top end); participants could choose any integer (Figure S2). The isiXhosa term used for the English ‘distress’ uses a metaphor that translates as “the weight you are feeling on your shoulders”, and is interpreted as referencing metaphorical (emotional) pressure or strain.

#### Pain

To assess pain, questions were modified from the Brief Pain Inventory (BPI) (50) (Figure S1), which is valid for isiXhosa-speaking adults living with HIV and pain (51). Questions addressed pain intensity, all sites of pain, and site of worst pain, each asked separately for pain in the past week (data presented here) and pain on most days for the past 3 months (not reported here).

### Data analysis

Data were deidentified and analysed using R version 4.4.0 (52) and RStudio version 2024-04-24 (53). Packages used are listed in the supplementary file.

#### Data pre-processing

We allocated study weeks to each data point, with the first study week starting on 16 March 2021. All data from seven participants were removed due to ineligibility for the parent study: six had provided inconsistent pain status responses between parent study screening and the baseline assessment; one had been pregnant. Figure 2 shows the inclusion and exclusion of participants and responses for each step in the study. To assess whether data missingness was informative, we first used linear regressions to determine whether a participant’s response rate was associated with their median rating of distress or pain. Second, we compared distress and pain reports between those who did vs did not contribute data to the time-lagged analyses.

**Fig. 2.**
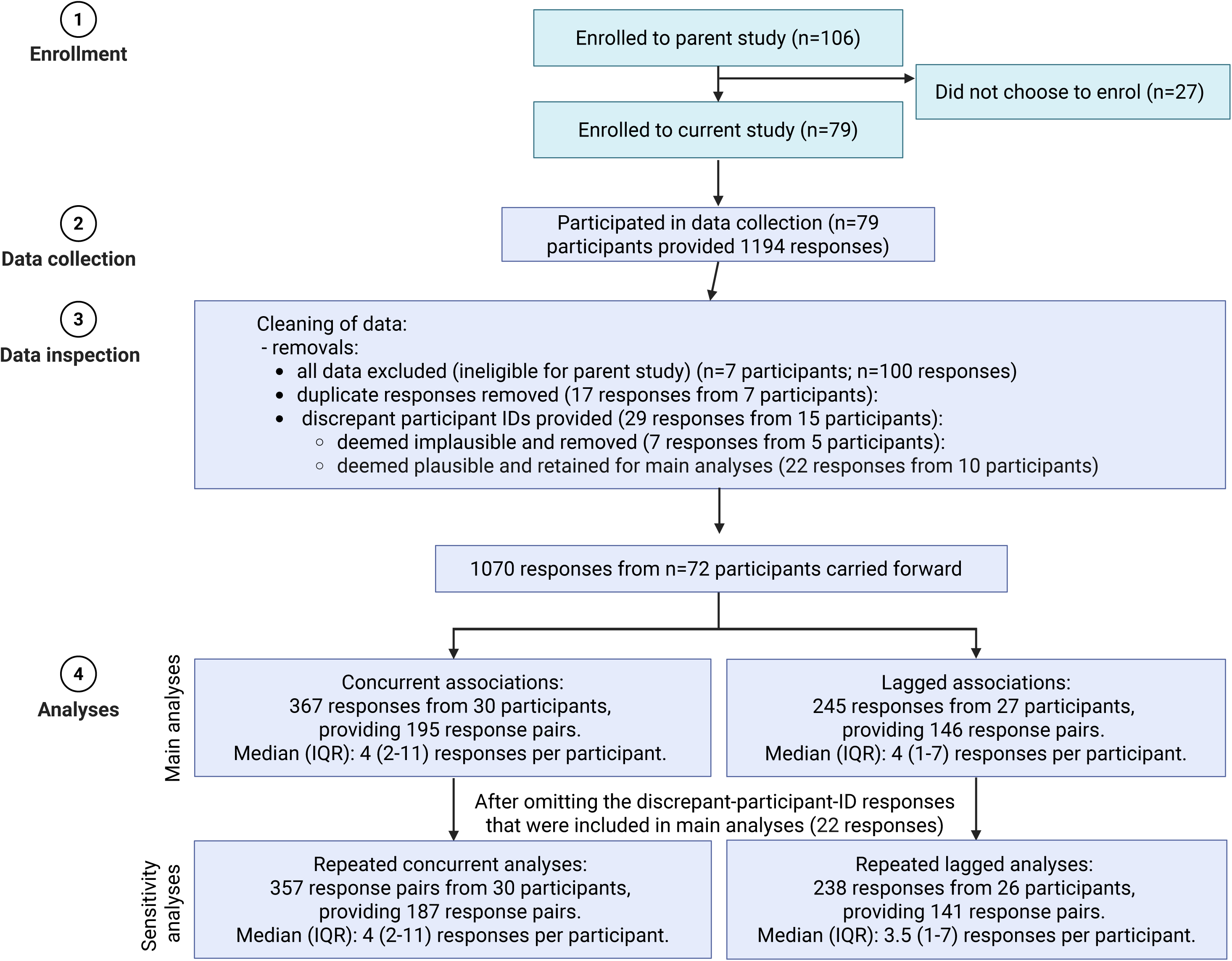
Flow of participants and responses through the study and analysis.

Seven participants had completed the survey more than once within a single week, together providing 17 duplicate responses; these responses were removed. In 29 responses from 15 participants, there was a mismatch between the unique app-generated user-ID and the self-reported participant study identifier (PID). These responses were closely inspected to ascertain whether the ID discrepancy could be due to innocent error (e.g. typing 333 instead of 033) and whether the content of the response with the ID mismatch was consistent with the content of responses preceding and following that response (e.g. consistently no pain, or pain site reported at another response from that participant). We made subjective judgements to remove 7 of the 29 responses with ID mismatch (e.g. ID 363 given instead of 024) (from 5 participants) and retain 22 (from 10 participants) for likely typing error (e.g. 030 given instead of 038) and consistency with previous responses (e.g. no pain, no pain, no pain). The 22 retained responses were excluded in sensitivity analyses that re-ran the unstandardised cumulative link mixed regression models from the main analyses, and these results are reported after the main analyses.

#### Statistical analysis

Variables are summarised with median and interquartile range (numerical) or frequencies and proportions (categorical). We visualised distress and pain ratings to observe distributions and individual-level fluctuations in ratings.

### Concurrent associations

To estimate the relationship between concurrent reports of distress and worst/average pain, we used four cumulative link mixed regression models (CLMMs). These models allow for ordinal data and assess the cumulative probability that higher ratings in the independent variable are statistically associated with higher ratings in the dependent variable. For two, distress was the dependent variable, and worst/average pain was the independent variable; for the other two, distress was the independent variable and worst/average pain was the dependent variable. We allowed random intercepts for individuals. Given the lack of nominal test available in R for CLMMs, we graphically assessed the proportional odds assumption (consistent relationship between predictor and outcome across all cumulative cut-points of the ordinal outcome). For each cumulative cut-point (0 vs 1–10, 0–1 vs 2–10, etc.), we fitted separate binary logistic regression models, plotted Odds Ratios (ORs) and 95% confidence intervals (95%CI), and deemed the assumption met if ORs were relatively consistent across cut-points. To test for differences in model estimates due to varying distributions of distress and pain intensity data, we computed unstandardized and standardized CLMMs.

### Time-lagged associations

One-week-time-lagged distress and pain scores were identified for every study week, for each respondent (n=72). Where a response was neither preceded nor followed by another response in an immediately consecutive study week, that response was excluded and represented as missing. The CLMMs’ structures were identical to the concurrent models except that dependent variable data were from the week after the independent variable.

As the proportional odds assumption was met for all the standardized and unstandardized CLMMs (Figures S11-14; S17-S20), we present only the main effects as a common odds ratio (OR) across all outcome cut-points. We describe unstandardised CLMM results in the text but report standardised CLMM results in Tables S4 and S6-S8.

We used Holm-Bonferroni corrections to control Type 1 error when testing multiple hypotheses using the same predictor, requiring 5 corrections in total. That is, 1 for the analysis of whether data missingness was informative; 1 for each of the 2 standardized; 1 for each of the 2 unstandardized models (main analysis and sensitivity analysis) with distress as predictor of worst/average pain intensity. Regression results are presented as odds ratios (OR) with 95% confidence intervals (CI’s); p-values are reported with the corrected threshold of 0.05.

## Results

This study analysed data from 72 participants, with median age of 43 (37–51) years. Of these 72 participants, 33 (46%) had initially reported persistent pain at enrolment to the parent study; the other 39 (54%) had initially reported no pain. There was a higher proportion of females (n=51; 71%) than males (n=21; 29%). The participants had very low income: mean monthly household income at enrolment to the parent study was ZAR3 508 (SD 2 935) (US$238), median household size was 3 (2, 4) people; 53% (n=38) lived in shack/informal housing, and 63% (n=45) were unemployed (Table S1).

Engagement with the questionnaires on the app was initially poor (see Figure S4). However, this improved with better staff support and incentivisation from late August 2021, tapered later, and improved again with a raffle incentive in March 2022. Response rate changes did not coincide with obvious changes in key study outcomes at the group level (Figure S5).

The distribution of responses over the 49 study weeks varied over time, with more responses occurring after 20 weeks, which was when study mobile phones were first distributed to participants (Figure S6a). There was high endorsement of ratings of 0 for distress (Figure S6b) and of ratings of 10 for pain intensity (Figures S6c and Sd).

Figure 2 shows the number of responses, response pairs, and participants represented in the dataset available for each step in the analysis. It also reports the median (IQR) number of responses per participant.

### Appraisal of data missingness

The two linear regressions showed that, on average, participants with higher median ratings of ‘worst pain’ had provided less than one more response in the study (beta-coefficient [95%CI] of 0.83 [0.12-1.54], p=0.023), whereas median distress rating was unrelated to the number of responses provided (Table S2). There were no other indications that data missingness was informative (see supplementary data for full reporting).

### Individual-level fluctuations in pain and distress

Figure S10 shows that there were some co-fluctuations in individuals’ pain and distress ratings over time, with considerable inter-individual variability.

### Concurrent associations

Worst pain and average pain statistically predicted distress in the same week (Table S3; Figure S15), and vice versa (Table S5; Figure S16). On average, a 1-point-higher rating of worst or average pain was associated with a 2.08 (1.77 – 2.43, p<0.001) and a 2.08 (1.79 – 2.42, p<0.001) increased odds of a higher distress score in the same week, respectively. A 1-point-higher distress rating was associated with a 90% (OR [95% CI]: 1.91 [1.64-2.22], p<0.001) or 85% (OR [95% CI]: 1.85 [1.61-2.13], p<0.001) greater odds of a higher worst or average pain score in the same week, respectively.

### Time-lagged associations

Worst pain and average pain predicted distress in the next week (Table 1; Figure 3). A 1-point-higher rating of worst pain or average pain was associated with a 42% (OR [95% CI]: 1.42 [1.17-1.71], p<0.001) or 43% (OR [95% CI]: 1.43 [1.20-1.71], p<0.001) greater odds of a higher distress score in the next week, respectively.

**Fig. 3.**
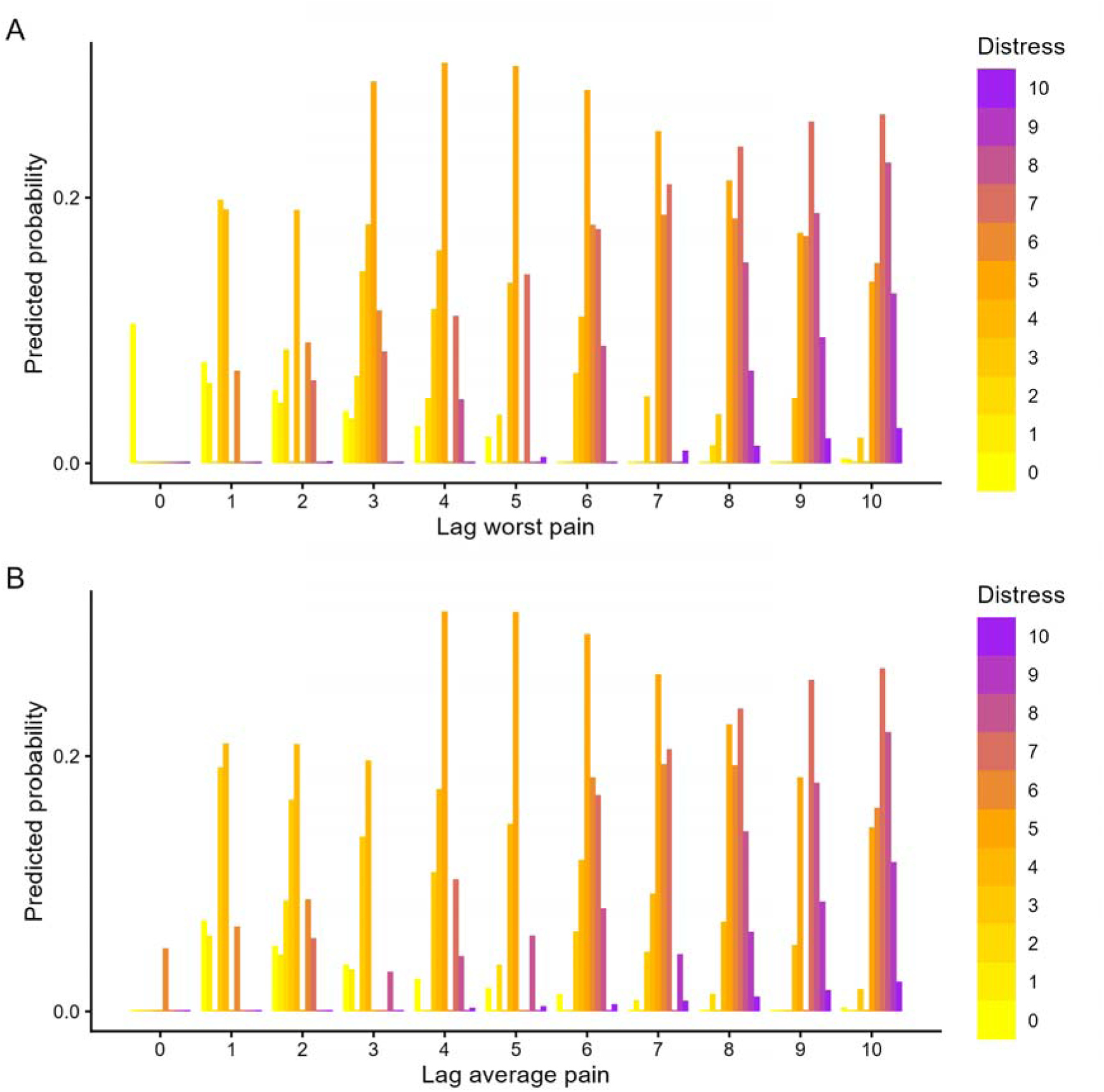
Association of each level of distress with each level of a) worst pain intensity or b) average pain intensity in the preceding week. Ratings in the lower range of the pain scale most strongly predicted distress of 0, 3, or 4/10 (yellow bars); ratings in the middle range of the pain scale most strongly predicted distress of 5/10 (orange bars), and ratings in the upper range of the pain scale most strongly predicted distress of 7/10 (purple bars), in the next week.

**Table 1.** The main effects of the unstandardized unadjusted cumulative link mixed models for worst/average pain intensity (as the independent variables) and distress in the next week (as the dependent variable).

|  | **Worst pain as predictor** |  |  | **Average pain as predictor** |  |  |
|---|---|---|---|---|---|---|
| *Predictors* | *Odds Ratio* | *CI* | *p* | *Odds Ratio* | *CI* | *p* |
| Lag worst pain | 1.42 | 1.17 – 1.71 | **<0.001** |  |  |  |
| Lag average pain |  |  |  | 1.43 | 1.20 – 1.71 | **<0.001** |
| **Random Effects** |  |  |  |  |  |  |
| σ<sup>2</sup> | 3.29 |  |  | 3.29 |  |  |
| τ<sub>00</sub> | 12.89<sub>blinded_upi_id</sub> |  |  | 13.58<sub>blinded_upi_id</sub> |  |  |
| ICC | 0.80 |  |  | 0.80 |  |  |
| N | 45<sub>blinded_upi_id</sub> |  |  | 45<sub>blinded_upi_id</sub> |  |  |
| Observations | 245 |  |  | 244 |  |  |
| Marginal R<sup>2</sup> / Conditional R<sup>2</sup> | 0.049 / 0.807 |  |  | 0.049 / 0.815 |  |  |

Distress predicted worst pain in the next week, but not average pain (Table 2; Figure 4). On average, a 1-point-higher rating of distress was statistically significantly associated with 25% (OR [95% CI]: 1.25 [1.07-1.46], p=0.023) greater odds of a higher rating of worst pain intensity, in the next week.

**Fig. 4.**
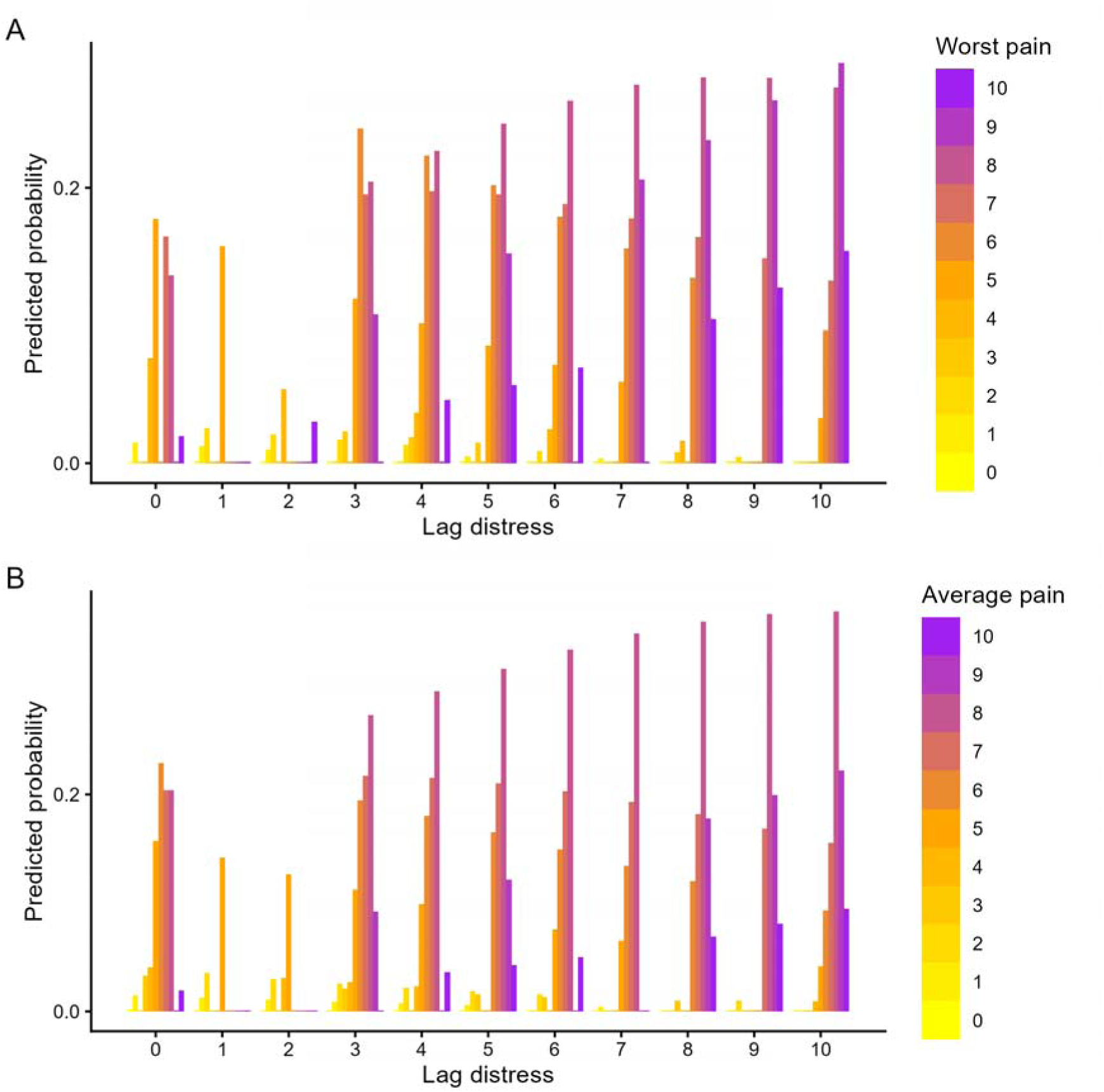
Association each level of distress in the past week with each level of a) worst pain intensity (statistically significant) and b) average pain intensity (not statistically significant) in the next week. Distress ratings of less than 3 most strongly predicted worst pain of 4, 5, 7, or 8/10; distress ratings of 3-6 most strongly predicted worst pain of 6, 7, or 8/10, and distress ratings of 7-10 most strongly predicted worst pain of 8 or 9/10.

**Table 2.** The main effects of the unstandardized unadjusted cumulative link mixed models for distress (as the independent variable), and worst or average pain intensity in the next week (as the dependent variables).

|  | **Worst pain as outcome** |  |  | **Average pain model as outcome** |  |  |
|---|---|---|---|---|---|---|
| *Predictors* | *Odds Ratio* | *CI* | *p* | *Odds Ratio* | *CI* | *p* |
| Lag distress | 1.25 | 1.07 – 1.46 | **0.023** | 1.19 | 1.01 – 1.40 | 0.152 |
| **Random Effects** |  |  |  |  |  |  |
| σ<sup>2</sup> | 3.29 |  |  | 3.29 |  |  |
| τ<sub>00</sub> | 7.61<sub>blinded_upi_id</sub> |  |  | 5.88<sub>blinded_upi_id</sub> |  |  |
| ICC | 0.70 |  |  | 0.64 |  |  |
| N | 47<sub>blinded_upi_id</sub> |  |  | 47<sub>blinded_upi_id</sub> |  |  |
| Observations | 255 |  |  | 255 |  |  |
| Marginal R<sup>2</sup> / Conditional R<sup>2</sup> | 0.043 / 0.711 |  |  | 0.031 / 0.653 |  |  |

#### Sensitivity analyses

Repeating the CLMMs without the 22 responses with discrepant participant identifiers yielded minimal changes in odds ratios and 95% confidence intervals (Tables S9-S12).

## Discussion

This study assessed week-to-week fluctuations in emotional distress and pain, and whether distress preceded changes in pain or pain preceded changes in distress, in people with HIV. Distress and pain intensity ratings were positively associated within the same time point. Higher worst and average pain temporally preceded higher distress the next week. Higher distress was associated with higher worst pain, but not with average pain, the next week.

Our findings replicate the bidirectional relationship between distress and pain that other studies have demonstrated using longer time periods (36, 54), and we confirmed that this relationship held true with one week’s delay and in people with HIV in this low-income context. It is interesting, given the load of socio-economic stressors poised to exacerbate general distress in people with HIV, that we did not find a more consistent association between distress and subsequent pain than vice versa. We speculate that this could be attributed to concern about pain. Ongoing diagnostic uncertainty is a common concern among people with HIV and pain (including in this sample (55)), as is pain-related worry (41, 55). Although our observational data cannot shed light on causal relationships, we speculate that diagnostic uncertainty could increase the potential for pain exacerbations to initiate or increase emotional distress (56, 57), particularly if increases in pain are interpreted as signs of progressing HIV disease or other threatening illness (55, 58, 59). Future work in this population should consider examining pain-focused distress, such as worrying/‘catastrophising’, alongside general distress. Designs that test causative hypotheses could also tease apart shared and overlapping mechanisms that may underpin relationships between general distress, pain, and pain-focused distress. Given that pain exacerbations may be especially distressing for people with fewer skills to manage their pain, it would be helpful to examine whether training people with HIV to manage their pain reduces the impact of pain on distress. Even without mechanistic clarity, people with HIV and pain are likely to benefit from training in self-management of pain and distress (60, 61).

Our finding of no association between average pain and preceding distress contrasts with previous studies where greater distress preceded greater average pain (62, 63). However, those studies used fewer observations, more participants, and longer time intervals (3-12 months), whereas we studied week-to-week changes, placing our work in the space between these multi-month time frames and other, ‘experience sampling’ designs (e.g., 64, 65). Moreover, because there is no isiXhosa term for ‘average’ (66), the pain ‘on average’ captured here may differ conceptually from studies with other populations. Notably, however, English-speaking research participants also use idiosyncratic and non-arithmetic approaches to rating pain, including to rate ‘average’ pain, which some consider synonymous with ‘usual’ or ‘everyday’ pain (67, 68). Both studies with findings contrasting ours included participants with clinically diagnosed chronic pain, whereas our participants were people with self-reported chronic or recurrent pain. Lastly, it is plausible that distress has different nuances and implications for our participants than for the previously studied, North American populations. Many people with HIV and low income face factors that can exacerbate distress and/or pain, and which fall outside their control due to pressures on livelihoods and low agency in employment situations (16, 43). The current study epitomised this poverty-related vulnerability alongside minimal formal support: some parent study participants reported considerable distress, but only 2% had accessed formal mental health care (69) although qualitative interviews indicated notable use of informal social support for distress and pain (55).

This study has several strengths. Participants and data collection staff were blinded to the research aims and hypotheses. The work addressed the pain-distress relationship in people with HIV, in a low-income South African setting, offering insights that may also have relevance to the many people with HIV who live in similarly low-resource contexts. Data were reported remotely from participants’ daily environments, reducing potential reporting bias associated with research environments. Deep characterisation of this sample (55, 69) enabled hypothesis generation and enhanced the utility of the findings for ongoing research and clinical application.

This work also has key limitations. Our sampling strategy excluded anyone who reported acute pain. Although our data indicate that some participants had neither persistent pain nor no pain by the time they provided data for the current study, our sample likely under-represents people with short-lived or intermittent pain. Indeed, the pattern of missing data suggests that our dataset is biased towards people with more pain, but the magnitude of that bias is unclear. Nonetheless, we cannot exclude the possibility that our results may be more applicable to those with more severe pain than to those with milder pain. The prospective relationships observed do not indicate causality; experimental manipulation (e.g. intervention to reduce either distress or pain) with frequent assessment of both would better support causal inference (70, 71). Although cross-lagged panel analysis would be preferable for assessing temporal relationships, this was not supported by the current dataset, which is better suited to ordinal regression (72, 73),. As noted above, the findings for ‘average pain’ should be interpreted with care. Similarly, the isiXhosa term used for ‘distress’ may place more emphasis on emotional aspects than the English term, although our experience is that local lay people use the English term ‘distress’ for emotional concerns. Formal testing of the face validity of the visual-numerical distress scale in this population would lay a stronger foundation for future local research needing brief measures. In our data, distress followed a bimodal distribution, whereas pain intensity was negatively skewed, resulting in lower variance in distress and potentially reduced regression accuracy. However, standardization did not change the strength or direction of associations. Finally, our data were collected within two years after the most stringent COVID-19 pandemic lockdowns, which were associated with increased depression in South Africa (74, 75) and we cannot rule out that this influenced our findings.

## Conclusion

This study found co-fluctuations between emotional distress and pain, a bidirectional relationship between distress and later worst pain, and a unidirectional relationship between average pain and later distress, in people with HIV. Building on these findings, future work should examine pain-focused constructs (e.g. pain catastrophizing and diagnostic uncertainty), to further focus interventions. Although causality could not be established, the observed bidirectional relationships between pain and distress, consistent with other studies, provides a compelling argument that clinical identification of pain or distress should prompt assessment for the other [49] to ensure that neither is missed if present, and that it could be helpful to develop and test clinical interventions that aim to concurrently address both pain and distress, for people with HIV.

## Data Availability

The de-identified data are available for selective sharing, subject to review of individual data requests, the use of secure computer platforms, formal use agreements, and compliance with the institutional
human research ethics policies. Data use is limited to research purposes, on secure computer servers. The principal investigator (VJM) is the contact person for requests to share data.

## Acknowledgements

VJM conceived the study. VJM, PK, JJ, RE, RP, and AW contributed to the design of the study. GA led the data analysis, interpretation of results and write-up of the manuscript with oversight from VJM. PK also supported data analysis. All authors read, commented on, and endorsed the final version of the manuscript for submission.

The authors thank Nomvula Mdwaba and Ncumisa Msolo for their data collection support.

This work was funded by NIH award K43TW011442 (VJM) and NRF South Africa Incentive Funding for Rated Researchers 119238 (AW). RRE was supported by NIH award K24 NS126570.

## References

1. Aaron RV, Ravyts SG, Carnahan ND, Bhattiprolu K, Harte N, McCaulley CC, et al. Prevalence of Depression and Anxiety Among Adults With Chronic Pain: A Systematic Review and Meta-Analysis. JAMA Netw Open. 2025;8(3):e250268.

2. Brooks JM, Umucu E, Huck GE, Fortuna K, Sánchez J, Chiu C, et al. Sociodemographic characteristics, health conditions, and functional impairment among older adults with serious mental illness reporting moderate-to-severe pain. Psychiatr Rehabil J. 2018;41(3):224–33.

3. Crombez G, Scott W, De Paepe AL. Knowing What We Are Talking About: The Case of Pain Catastrophizing. J Pain. 2024;25(3):591–4.

4. Vlaeyen JWS, Linton SJ. Fear-avoidance model of chronic musculoskeletal pain: 12 years on. Pain. 2012;153(6):1144–7.

5. Vlaeyen JWS, Linton SJ. Fear-avoidance and its consequences in chronic musculoskeletal pain: a state of the art. Pain. 2000;85(3):317–32.

6. Edwards RR, Bingham CO, 3rd, Bathon J, Haythornthwaite JA. Catastrophizing and pain in arthritis, fibromyalgia, and other rheumatic diseases. Arthritis Rheum. 2006;55(2):325–32.

7. Rogers AH, Farris SG. A meta-analysis of the associations of elements of the fear-avoidance model of chronic pain with negative affect, depression, anxiety, pain-related disability and pain intensity. Eur J Pain. 2022;26(8):1611–35.

8. Theunissen M, Peters ML, Bruce J, Gramke HF, Marcus MA. Preoperative anxiety and catastrophizing: a systematic review and meta-analysis of the association with chronic postsurgical pain. The Clinical journal of pain. 2012;28(9):819–41.

9. George SZ, Dannecker EA, Robinson ME. Fear of pain, not pain catastrophizing, predicts acute pain intensity, but neither factor predicts tolerance or blood pressure reactivity: an experimental investigation in pain-free individuals. Eur J Pain. 2006;10(5):457–65.

10. Zhao X, Boersma K, Gerdle B, Molander P, Hesser H. Fear network and pain extent: Interplays among psychological constructs related to the fear-avoidance model. J Psychosom Res. 2023;167:111176.

11. Nieto R, Miró J, Huguet A. Pain-related fear of movement and catastrophizing in whiplash-associated disorders. Rehabil Psychol. 2013;58(4):361–8.

12. Li J, Cui Y, Jia Q, Ouyang A, Hua Y. Pain Intensity and Pain Catastrophizing Among Patients with Chronic Pain: The Mediating Effect of Self-Efficacy. J Pain Res. 2025;18:1361–73.

13. Wood TJ, Thornley P, Petruccelli D, Kabali C, Winemaker M, de Beer J. Preoperative Predictors of Pain Catastrophizing, Anxiety, and Depression in Patients Undergoing Total Joint Arthroplasty. J Arthroplasty. 2016;31(12):2750–6.

14. Campbell CM, McCauley L, Bounds SC, Mathur VA, Conn L, Simango M, et al. Changes in pain catastrophizing predict later changes in fibromyalgia clinical and experimental pain report: cross-lagged panel analyses of dispositional and situational catastrophizing. Arthritis Res Ther. 2012;14(5):R231.

15. Crawford A, Muere A, Tripp DA, Nickel JC, Doiron RC, Moldwin R, et al. The chicken or the egg: Longitudinal changes in pain and catastrophizing in women with interstitial cystitis/bladder pain syndrome. Can Urol Assoc J. 2021;15(10):326–31.

16. Scott W, Arkuter C, Kioskli K, Kemp H, McCracken LM, Rice ASC, et al. Psychosocial factors associated with persistent pain in people with HIV: a systematic review with meta-analysis. Pain. 2018;159(12):2461–76.

17. Van Loey NE, Klein-König I, de Jong AEE, Hofland HWC, Vandermeulen E, Engelhard IM. Catastrophizing, pain and traumatic stress symptoms following burns: A prospective study. Eur J Pain. 2018;22(6):1151–9.

18. Kroska EB. A meta-analysis of fear-avoidance and pain intensity: The paradox of chronic pain. Scand J Pain. 2016;13:43–58.

19. Jakobsen MD, Vinstrup J, Andersen LL. Work-Related Fear-Avoidance Beliefs and Risk of Low-Back Pain: Prospective Cohort Study Among Healthcare Workers. J Occup Rehabil. 2025;35(3):547–55.

20. Zhao Z, Li J, Zhang R, Feng Y, He Y, Sun Z. The prognostic value of fear-avoidance beliefs on postoperative pain and dysfunction for lumbar degenerative disk disease: a meta-analysis. Int J Rehabil Res. 2023;46(1):3–13.

21. Asiri F, Reddy RS, Tedla JS, MA AL, Alshahrani MS, Govindappa SC, et al. Kinesiophobia and its correlations with pain, proprioception, and functional performance among individuals with chronic neck pain. PLoS One. 2021;16(7):e0254262.

22. Bakırhan S, Unver B, Elibol N, Karatosun V. Fear of movement and other associated factors in older patients with total knee arthroplasty. Ir J Med Sci. 2023;192(5):2217–22.

23. Kolodziejczak-Krupp K, Wilhelm LO, Diering LE, Zipper V, Maas J, Schäfer T, et al. Psychological risk factors and resources for low back pain intensity and back health in daily life: An ecological momentary assessment study. Appl Psychol Health Well Being. 2025;17(5):e70080.

24. Monticone M, Arippa F, Frigau L, Foti C, Ferrari S, Guicciardi M, et al. Model of Fear of Movement/(Re)injury Runs Clockwise From Catastrophizing: Evidence From a Sample of Outpatients With Chronic Nonspecific Low Back Pain. Am J Phys Med Rehabil. 2025;104(6):506–10.

25. Timmers I, Quaedflieg CWEM, Hsu C, Heathcote LC, Rovnaghi CR, Simons LE. The interaction between stress and chronic pain through the lens of threat learning. Neuroscience & Biobehavioral Reviews. 2019;107:641–55.

26. Gandhi R, Tsvetkov D, Dhottar H, Davey JR, Mahomed NN. Quantifying the pain experience in hip and knee osteoarthritis. Pain Res Manag. 2010;15(4):224–8.

27. Yuan Y, Schreiber K, Flowers KM, Edwards R, Azizoddin D, Ashcraft L, et al. The relationship between emotion regulation and pain catastrophizing in patients with chronic pain. Pain Med. 2024;25(7):468–77.

28. Pincus T, Burton AK, Vogel S, Field AP. A Systematic Review of Psychological Factors as Predictors of Chronicity/Disability in Prospective Cohorts of Low Back Pain. Spine. 2002;27(5):E109–E20.

29. Jackson T, Tian P, Wang Y, Iezzi T, Xie W. Toward Identifying Moderators of Associations Between Presurgery Emotional Distress and Postoperative Pain Outcomes: A Meta-Analysis of Longitudinal Studies. J Pain. 2016;17(8):874–88.

30. Larouche M, Brotto LA, Koenig NA, Lee T, Cundiff GW, Geoffrion R. Depression, Anxiety, and Pelvic Floor Symptoms Before and After Surgery for Pelvic Floor Dysfunction. Female Pelvic Med Reconstr Surg. 2020;26(1):67–72.

31. Brinkman N, Thomas JE, Teunis T, Ring D, Gwilym S, Jayakumar P. Recovery of Comfort and Capability After Upper Extremity Fracture Is Predominantly Associated With Mindset: A Longitudinal Cohort From the United Kingdom. J Orthop Trauma. 2024;38(10):557–65.

32. Lingard EA, Riddle DL. Impact of psychological distress on pain and function following knee arthroplasty. J Bone Joint Surg Am. 2007;89(6):1161–9.

33. Dunn LK, Durieux ME, Fernández LG, Tsang S, Smith-Straesser EE, Jhaveri HF, et al. Influence of catastrophizing, anxiety, and depression on in-hospital opioid consumption, pain, and quality of recovery after adult spine surgery. J Neurosurg Spine. 2018;28(1):119–26.

34. Mirzaie S, Oberoi MK, Huang KX, Caprini RM, Malapati SH, Dejam D, et al. Association of Patient-Reported Anxiety and Pain After Alveolar Bone Grafting. Cleft Palate Craniofac J. 2024;61(8):1336–43.

35. Sulen N, Šimurina T, Požgain I, Župčić M, Sorić T, Basioli Kasap E, et al. Effects of preoperative anxiety, depression and pain on quality of postoperative recovery and acute postoperative pain after radical prostatectomy: a prospective observational study. Acta Clin Croat. 2023;62(4):677–87.

36. Engel F, Stadnitski T, Stroe-Kunold E, Berens S, Schäfert R, Wild B. Temporal Relationships Between Abdominal Pain, Psychological Distress and Coping in Patients With IBS - A Time Series Approach. Front Psychiatry. 2022;13:768134.

37. Virgolino A, Costa J, Santos O, Pereira ME, Antunes R, Ambrósio S, et al. Lost in transition: a systematic review of the association between unemployment and mental health. J Ment Health. 2022;31(3):432–44.

38. Lund C, Breen A, Flisher AJ, Kakuma R, Corrigall J, Joska JA, et al. Poverty and common mental disorders in low and middle income countries: A systematic review. Soc Sci Med. 2010;71(3):517–28.

39. Lund C, Brooke-Sumner C, Baingana F, Baron EC, Breuer E, Chandra P, et al. Social determinants of mental disorders and the Sustainable Development Goals: a systematic review of reviews. Lancet Psychiatry. 2018;5(4):357–69.

40. Wiginton JM, Murray S, Kall M, Maksut JL, Augustinavicius J, Delpech V, et al. HIV-related Stigma and Discrimination in Health Care and Health-related Quality of Life among People Living with HIV in England and Wales: A Latent Class Analysis. Stigma Health. 2023;8(4):487–96.

41. Sipilä R, Kemp H, Harno H, Rice ASC, Kalso E. Health-related quality of life and pain interference in two patient cohorts with neuropathic pain: breast cancer survivors and HIV patients. Scand J Pain. 2021;21(3):512–21.

42. de Souza A, Caumo W, Calvetti PU, Lorenzoni RN, da Rosa GK, Lazzarotto AR, et al. Comparison of pain burden and psychological factors in Brazilian women living with HIV and chronic neuropathic or nociceptive pain: An exploratory study. PLoS One. 2018;13(5):e0196718.

43. Pham VA, Nguyen H, Krakauer EL, Harding R. “I Wish I Could Die So I Would Not Be in Pain”: A Qualitative Study of Palliative Care Needs Among People With Cancer or HIV/AIDS in Vietnam and Their Caregivers. J Pain Symptom Manage. 2021;62(2):364–72.

44. Merlin JS, Zinski A, Norton WE, Ritchie CS, Saag MS, Mugavero MJ, et al. A Conceptual Framework for Understanding Chronic Pain in Patients with HIV. Pain Practice. 2014;14(3):207–16.

45. Nguyen AL, Lake JE, Reid MC, Glasner S, Jenkins J, Candelario J, et al. Attitudes towards exercise among substance using older adults living with HIV and chronic pain. AIDS care. 2017;29(9):1149–52.

46. Brandt CP, Zvolensky MJ, Daumas SD, Grover KW, Gonzalez A. Pain-related anxiety in relation to anxiety and depression among persons living with HIV/AIDS. AIDS care. 2016;28(4):432–5.

47. Joseph V, Jones A, Canidate S, Mannes Z, Lu H, Ennis N, et al. Factors associated with current and severe pain among people living with HIV: results from a statewide sample. BMC public health. 2020;20(1):1424.

48. Adams LM, Volgenau KM, Regalario I, Hunt AD. Examining the relationships between pain symptoms and psychosocial functioning among women living with and at risk for human immunodeficiency virus using a cross-sectional psychological network analysis. Rehabil Psychol. 2025;70(3):320–31.

49. Madden VJ, Msolo N, Mqadi L, Lesosky M, Bedwell GJ, Hutchinson MR, et al. Study protocol: an observational study of distress, immune function and persistent pain in HIV. BMJ open. 2022;12(6).

50. Cleeland CS, Ryan KM. Pain assessment: Global use of the Brief Pain Inventory. Annals, Academy of Medicine, Singapore. 1994;23(2):129–38.

51. Parker R, Jelsma J, Stein DJ. Pain in amaXhosa Women Living With HIV/AIDS: Translation and Validation of the Brief Pain Inventory–Xhosa. Journal of Pain and Symptom Management. 2016;51(1):126–32.e2.

52. R Core Team. R: A Language and Environment for Statistical Computing. Vienna: Austria: R Foundation for Statistical Computing; 2022.

53. Posit team. RStudio: Integrated Development Environment for R. Boston, MA: Posit software, PBC; 2025.

54. Racine M, Moulin DE, Nielson WR, Morley-Forster PK, Lynch M, Clark AJ, et al. The reciprocal associations between catastrophizing and pain outcomes in patients being treated for neuropathic pain: a cross-lagged panel analysis study. Pain. 2016;157(9):1946–53.

55. Madden VJ, Court L, Gidana A, Knight L, Wadley A, Mqadi L, et al. Informal social support for chronic pain and distress in people with HIV: identifying targets for intervention. medRxiv. 2025:2025.11.07.25339650.

56. Martin S, Smith M, Wilson DA, Zadro JR, Ferreira GE, O’Keeffe M. Non-specific diagnostic labels for musculoskeletal conditions foster positive views about prognosis and non-invasive management but require clear explanation: a systematic review. J Physiother. 2025;71(3):167–78.

57. Serbic D, Evangeli M, Probyn K, Pincus T. Health-related guilt in chronic primary pain: A systematic review of evidence. Br J Health Psychol. 2022;27(1):67–95.

58. Mulaudzi M, Ratshinanga A, Mohale J, Ramoshai T, Evangeli M, Pincus T, et al. Experiences and impact of chronic pain in South Africans living in a rural area: a qualitative study. BMJ open. 2025;15(12):e103307.

59. Ownby KK, Dune LS. The processes by which persons with HIV-related peripheral neuropathy manage their symptoms: a qualitative study. J Pain Symptom Manage. 2007;34(1):48–59.

60. Ernstzen D, Keet J, Louw K-A, Park-Ross J, Pask L, Reardon C, et al. “So, you must understand that that group changed everything”: perspectives on a telehealth group intervention for individuals with chronic pain. BMC Musculoskeletal Disorders. 2022;23(1):538.

61. Merlin JS, Westfall AO, Long D, Davies S, Saag M, Demonte W, et al. A Randomized Pilot Trial of a Novel Behavioral Intervention for Chronic Pain Tailored to Individuals with HIV. AIDS and behavior. 2018;22(8):2733–42.

62. Dunbar MS, Rodriguez A, Edelen MO, Hays RD, Coulter ID, Siconolfi D, et al. Longitudinal Associations of PROMIS-29 Anxiety and Depression Symptoms With Low Back Pain Impact in a Sample of U.S. Military Service Members. Mil Med. 2023;188(3-4):e630–e6.

63. Sturgeon JA, Langford D, Tauben D, Sullivan M. Pain Intensity as a Lagging Indicator of Patient Improvement: Longitudinal Relationships With Sleep, Psychiatric Distress, and Function in Multidisciplinary Care. J Pain. 2021;22(3):313–21.

64. Bosman M, Vork L, Van Den Houte M, van Oudenhove L, Midenfjord I, Simren M, et al. Real-Time Abdominal Pain-Daily Life Trigger Associations in Irritable Bowel Syndrome: A Multicenter Experience Sampling Method Study. Clin Gastroenterol Hepatol. 2026.

65. Kuper QTY, Willemen JE, Lousberg R, Severeijns RJM, van Amelsvoort T, Balthasar AJR. How Affective States Moderate the Relationship of Pain Catastrophizing With Pain Severity in Patients With Acute and Chronic Pain. Brain Behav. 2026;16(10):e71820.

66. Mphahlele NR, Mitchell D, Kamerman PR. Pain in ambulatory HIV-positive South Africans. Eur J Pain. 2012;16(3):447–58.

67. Robinson-Papp J, George MC, Dorfman D, Simpson DM. Barriers to Chronic Pain Measurement: A Qualitative Study of Patient Perspectives. Pain Med. 2015;16(7):1256–64.

68. de Williams AC, Davies HTO, Chadury Y. Simple pain rating scales hide complex idiosyncratic meanings. Pain. 2000;85(3):457–63.

69. Madden VJ, Mqadi L, Arendse G, Bedwell GJ, Msolo N, Lesosky M, et al. Provoked cytokine response is not associated with distress or induced secondary hyperalgesia in people with suppressed HIV. Pain. 2025.

70. Vlaeyen JWS, Wicksell RK, Simons LE, Gentili C, De TK, Tate RL, et al. From Boulder to Stockholm in 70 Years: Single Case Experimental Designs in Clinical Research. The Psychological Record. 2020;70(4):659–70.

71. Tanious R, Madden VJ, Sharma S, Caneiro JP, Kwan LJ, Wicksell RK, et al. Emerging clinical pain research in low- and middle-income settings: towards a person-centred approach. Pain. 2026;167(10):2240–7.

72. Falkenström F, Solomonov N, Rubel J. Using Time-Lagged Panel Data Analysis to Study Mechanisms of Change in Psychotherapy Research: Methodological Recommendations. Couns Psychother Res. 2020;20(3):435–41.

73. Yoon JY, Brown RL. Causal inference in cross-lagged panel analysis: a reciprocal causal relationship between cognitive function and depressive symptoms. Res Gerontol Nurs. 2014;7(4):152–8.

74. Casale D, and Shepherd D. The gendered effects of the Covid-19 crisis in South Africa: Evidence from NIDS-CRAM waves 1–5. Development Southern Africa. 2022;39(5):644–63.

75. Waterfield KC, Shah GH, Etheredge GD, Ikhile O. Consequences of COVID-19 crisis for persons with HIV: the impact of social determinants of health. BMC public health. 2021;21(1):299.

