## Supplementary file for "Temporal relationships between distress and pain in people living with HIV"

### Supplementary materials

### Methods

#### Procedures

From 30 August 2021, funding support allowed us to implement 4 changes: (1) a dedicated recruiter began work on the study, (2) each participant could borrow a study mobile phone (Mobicel FAME 16GB single sim) to facilitate participation and was informed that phone ownership would be transferred to them after the study if they responded to over >80% of questionnaires, (3) a questionnaire was added with questions on COVID-19 experiences (data not reported here), and (4) a ZAR10 (US$0.6) airtime voucher was sent to compensate participants who had completed both questionnaires, each week. Another strategy was implemented from 28 February to 28 March 2022, to address tapering response rates: participants who completed both weekly questionnaires were entered into a weekly raffle to win a ZAR100 (US$5.60) grocery store or airtime voucher. After study completion, participants were asked to return the loaned phones. At this time, the threshold for ownership transfer was revised to be more pragmatic; those with >70% response rate received phone ownership.

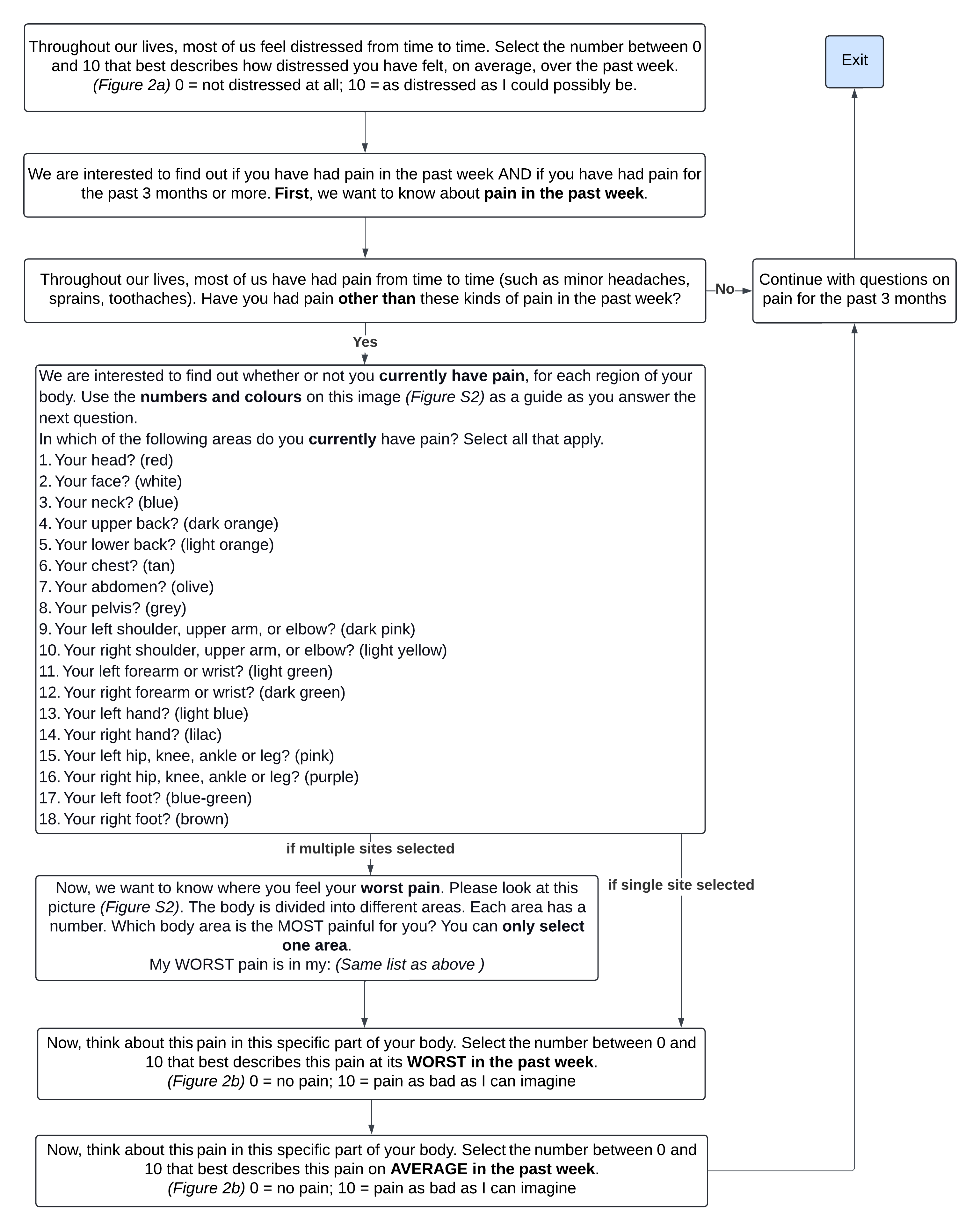
 **Figure S1.** Flow diagram with complete list of questions.

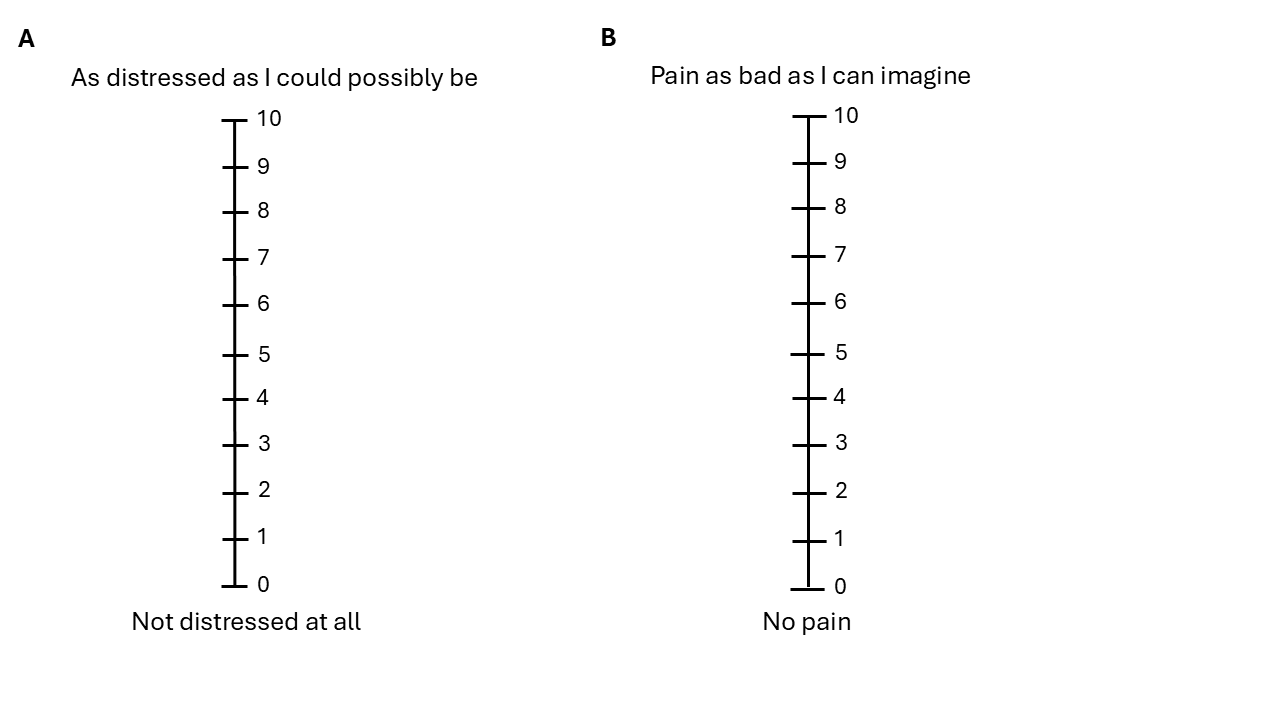

**Figure S2.** Visual analogue scale for a) distress and b) pain intensity.

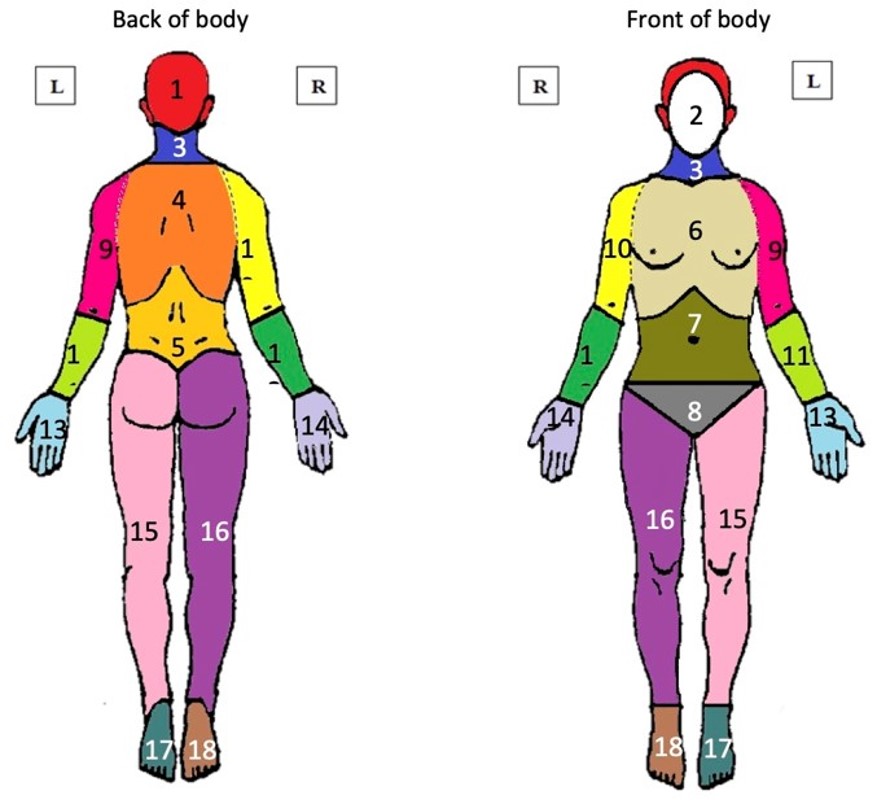

**Figure S3.** Site locations as described on body map.

#### Data analysis

We used packages knitr [89], dplyr [85], tidyverse [86], tidyr [82], readr [83], magrittr [5], flextable [32], gtsummary [73], gt [34], ggplot2 [81], patchwork [63], data.table [20], sjPlot [44], webshot [14], ordinal [15], MetBrewer [7], purrr [33], broom [68], sessioninfo [84], bibtex [29].

### Results

**Table S1.** Descriptive characteristics of sample of n=72 participants.

| **Characteristic** | **N = 72**^1^ |
| --- | --- |
| **Age (years)** | 43 (37, 51) |
| **Sex** |  |
| female | 51 (71%) |
| male | 21 (29%) |
| **Household income (Rand)** | 3,508 (2,935) |
| **Household size** | 3 (2, 4) |
| **Housing type** |  |
| formal housing | 34 (47%) |
| shack/informal housing | 38 (53%) |
| **Employment status** |  |
| unemployed | 45 (63%) |
| employed | 26 (37%) |
| missing | 1 |
| ^1^Median (Q1, Q3); n (%); Mean (SD) | |

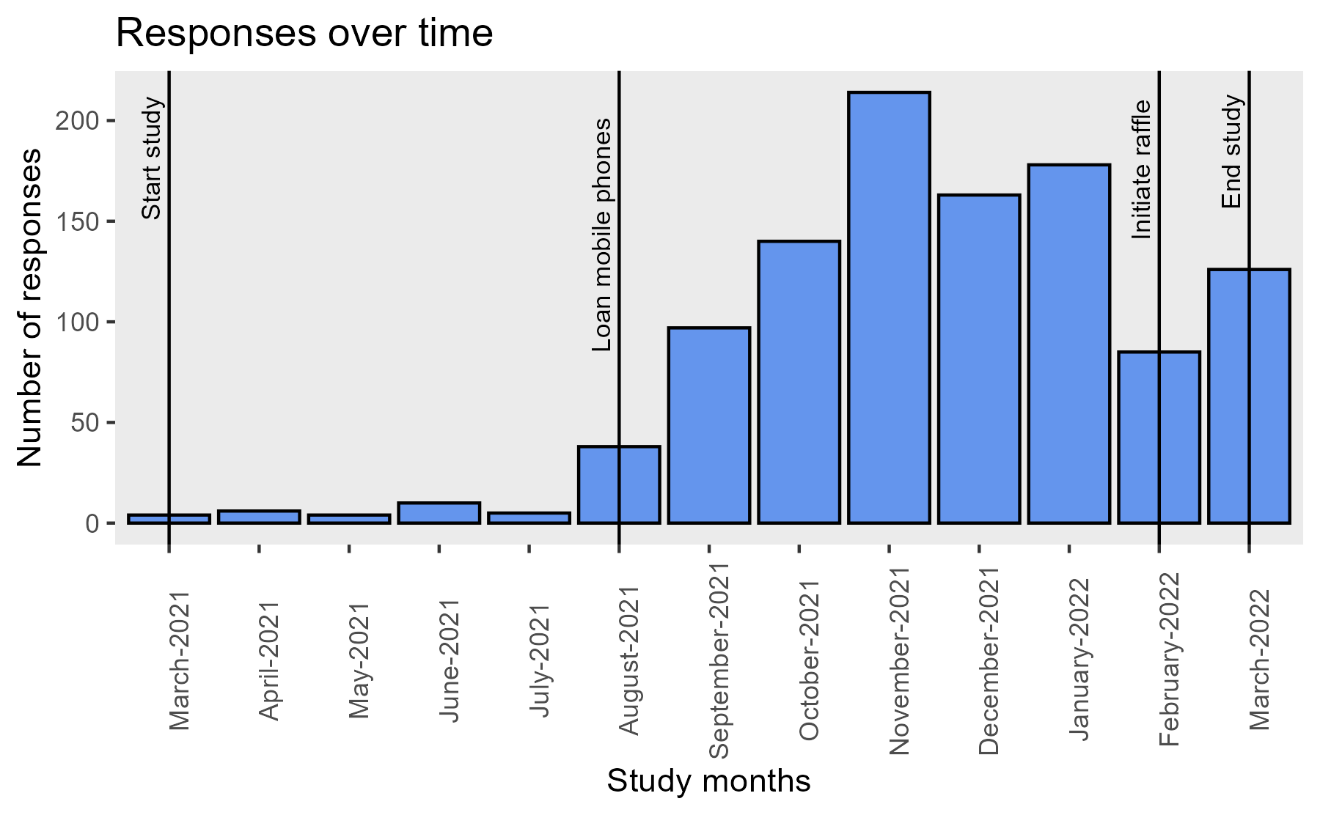
 **Figure S4.** Response rate and study timeline.

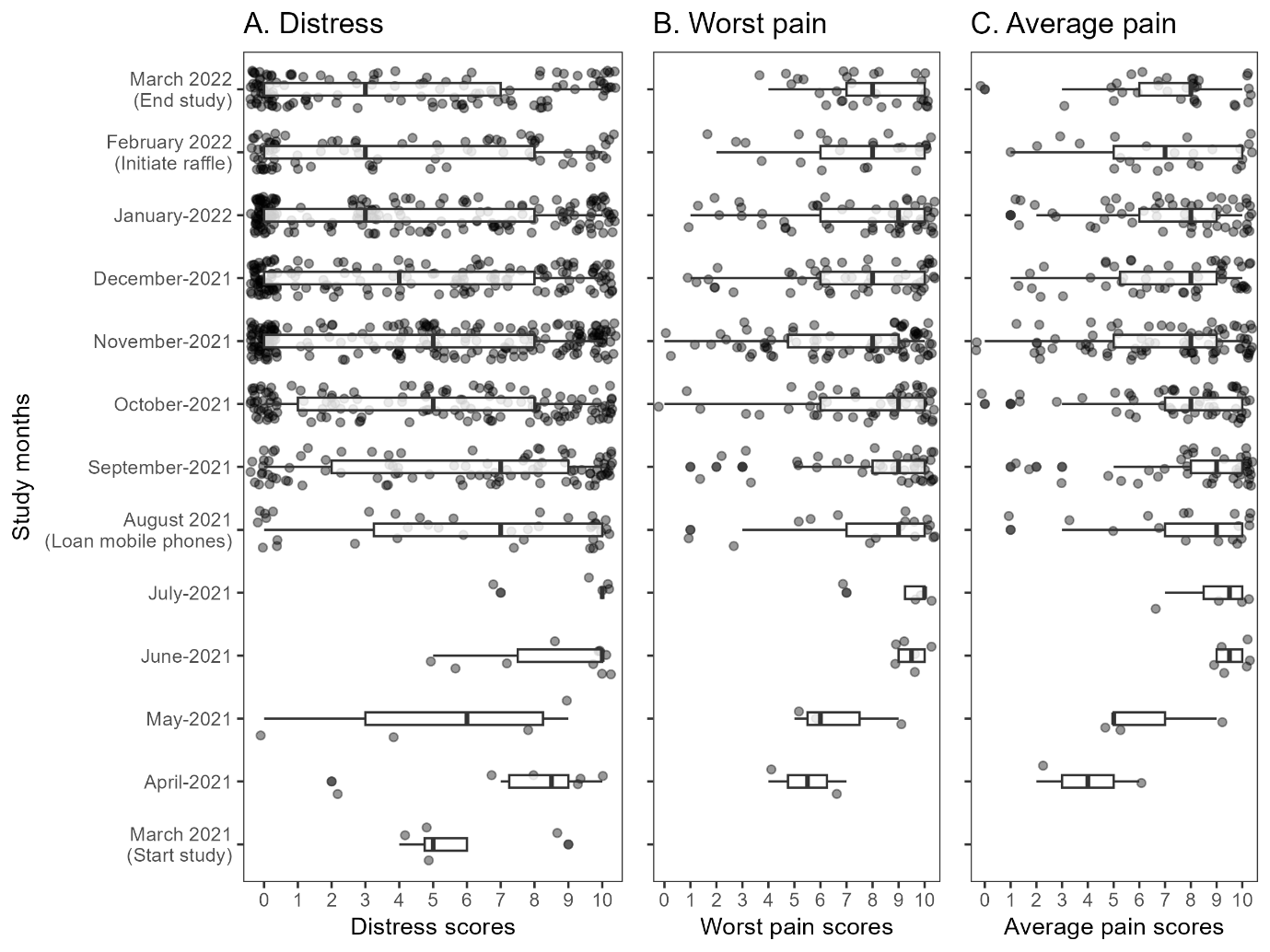
 **Figure S5.** Box-and-whisker plots to visually assess for a relationship between self-reported outcomes and incentivisation strategies (noted in y-axis labels). Outcomes plotted are: a) distress, b) worst pain severity and c) average pain severity and study timeline. Points show individual scores.

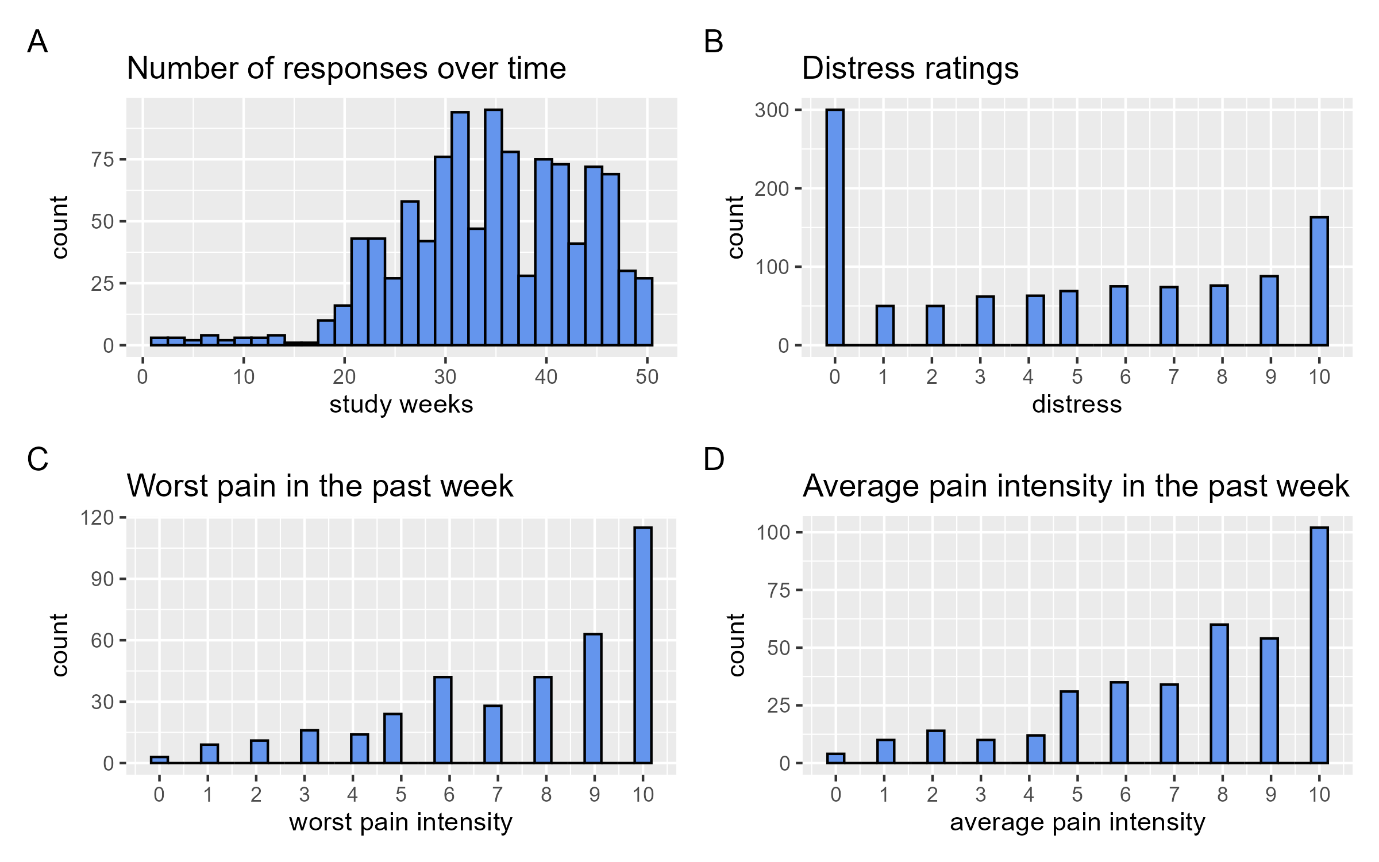
**Figure S6.** Distribution of the a) the number of responses over the study weeks, b) distress ratings, c) worst pain intensity ratings and d) average pain intensity ratings.

#### Appraisal of data missingness

| Table S2: Unadjusted linear regression models for number of participant responses (outcome) being predicted by (left model) distress or (right model) ‘worst pain’ intensity in the past week; p-values reported after Holm-bonferroni correction. | | | | | | |
| --- | --- | --- | --- | --- | --- | --- |
|  | **Distress as predictor** | | | **Worst pain as predictor** | | |
| *Predictors* | *Estimates* | *CI* | *p* | *Estimates* | *CI* | *p* |
| median distress | 0.49 | -0.11 – 1.10 | 0.109 |  |  |  |
| median worst pain |  |  |  | 0.83 | 0.12 – 1.54 | **0.023** |
| Observations | 57 | | | 57 | | |
| R^2^ / R^2^ adjusted | 0.046 / 0.029 | | | 0.090 / 0.074 | | |

Our visual comparison of within-participant median distress ratings between those who did vs did not contribute data to the lagged analyses showed no noteworthy difference (Figure S7).

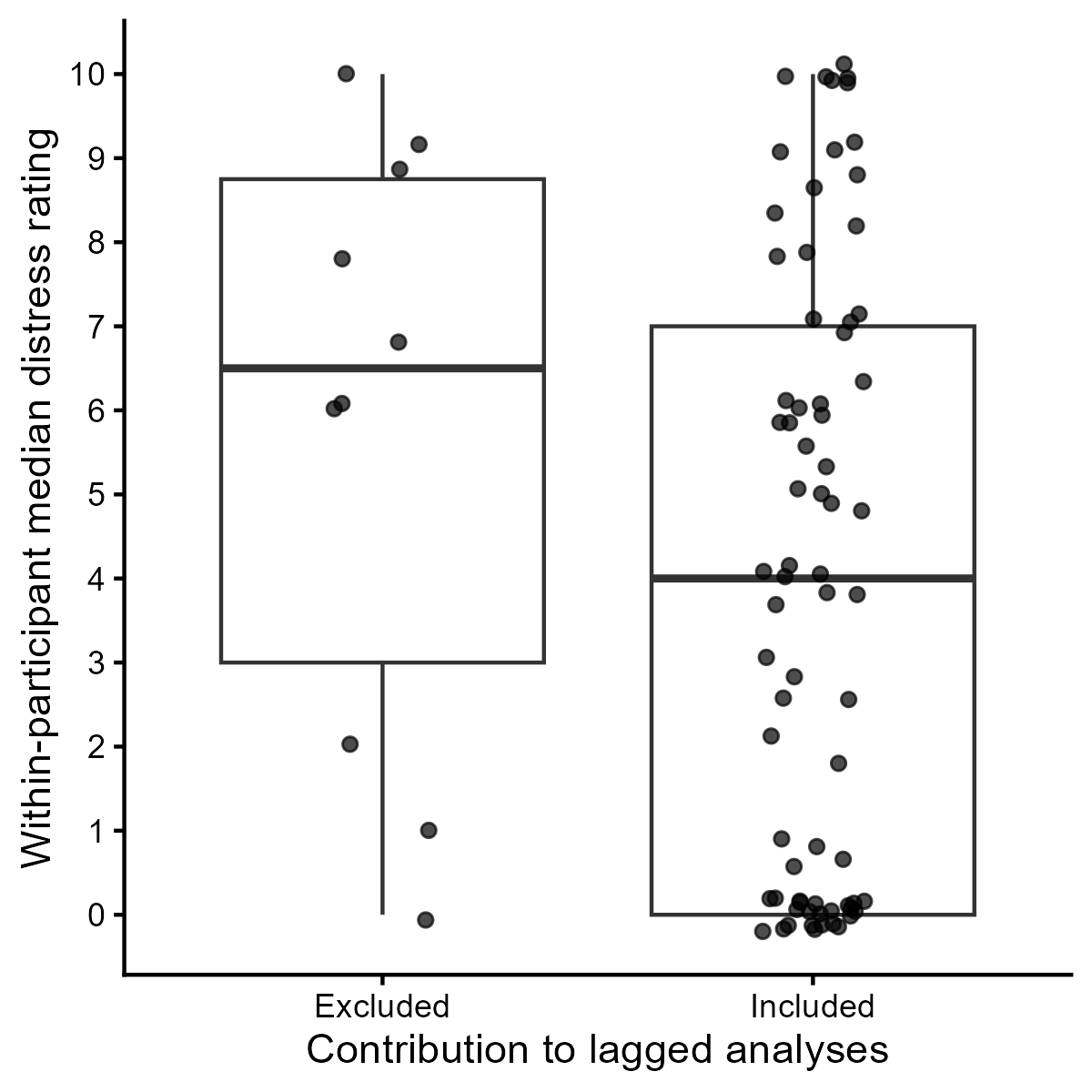

Figure S7: Median distress ratings were not notably different between those whose data did vs did not contribute the lagged analyses.

The lagged analyses included data from 50 participants with and 2 without pain, whereas all 10 participants whose 22 responses were omitted from the lagged analysis had reported pain at least once during the study (Figure S8).

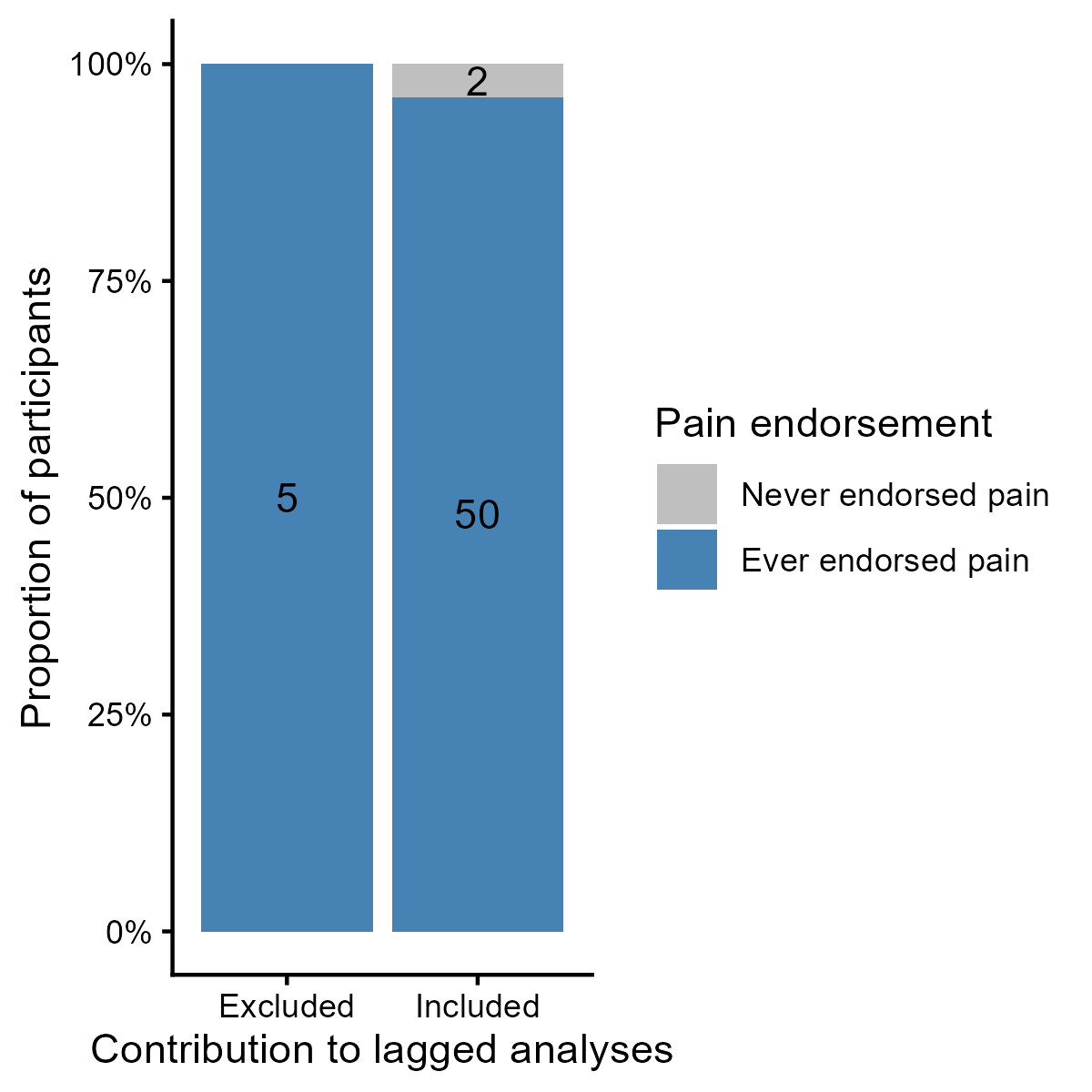

Figure S8: All 10 participants whose data did not contribute the lagged analyses had reported pain at some point during the study, whereas 50 of those whose data did contribute had endorsed pain.

Comparison of data from those who had reported pain at least once showed no noteworthy differences in within-participant median rating of ‘worst pain’ (Figure S9).

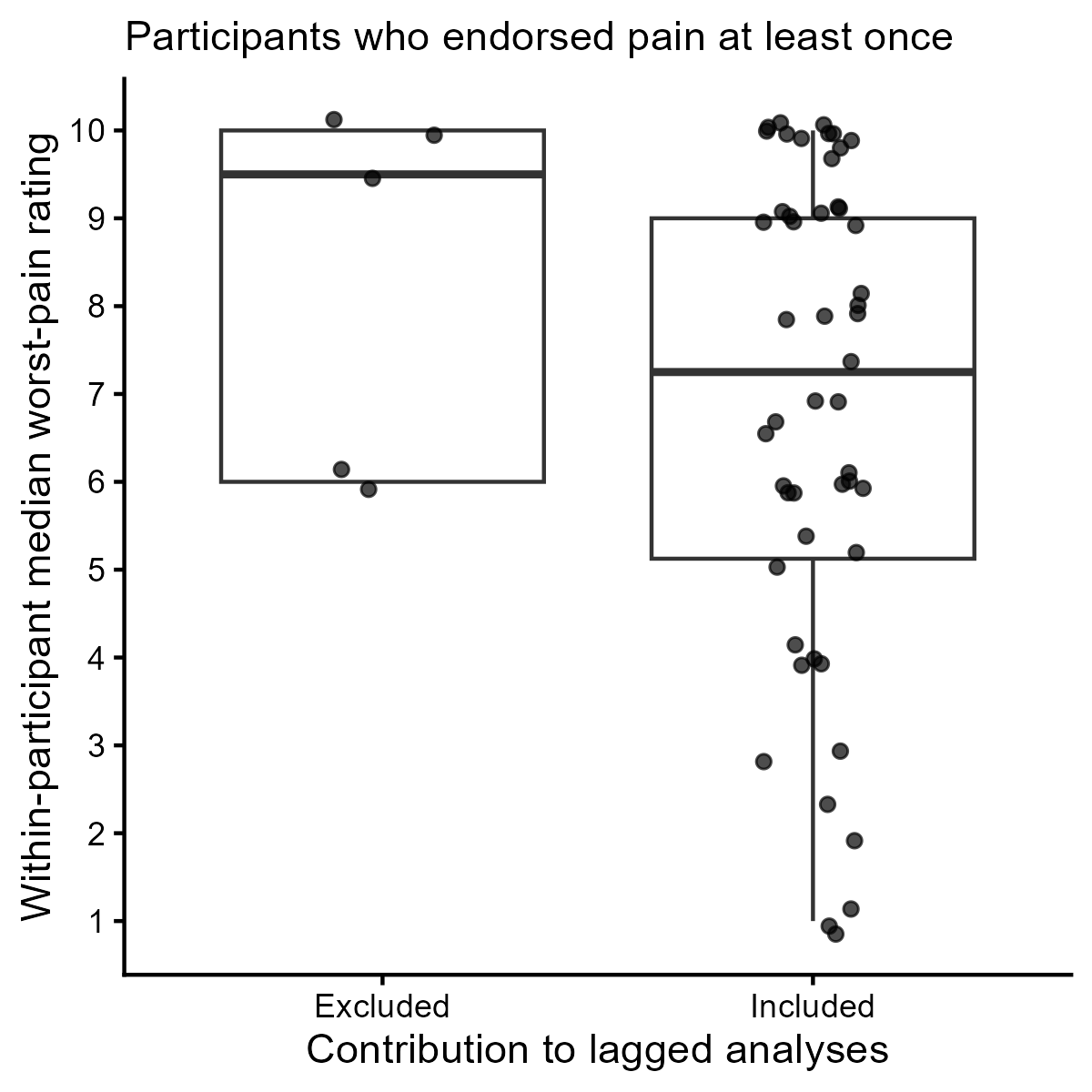

Figure S9: Of the participants who had reported pain at some point during the study, median ‘worst pain’ ratings were not notably different between those whose data did vs did not contribute the lagged analyses.

#### Individual-level fluctuations in pain and distress

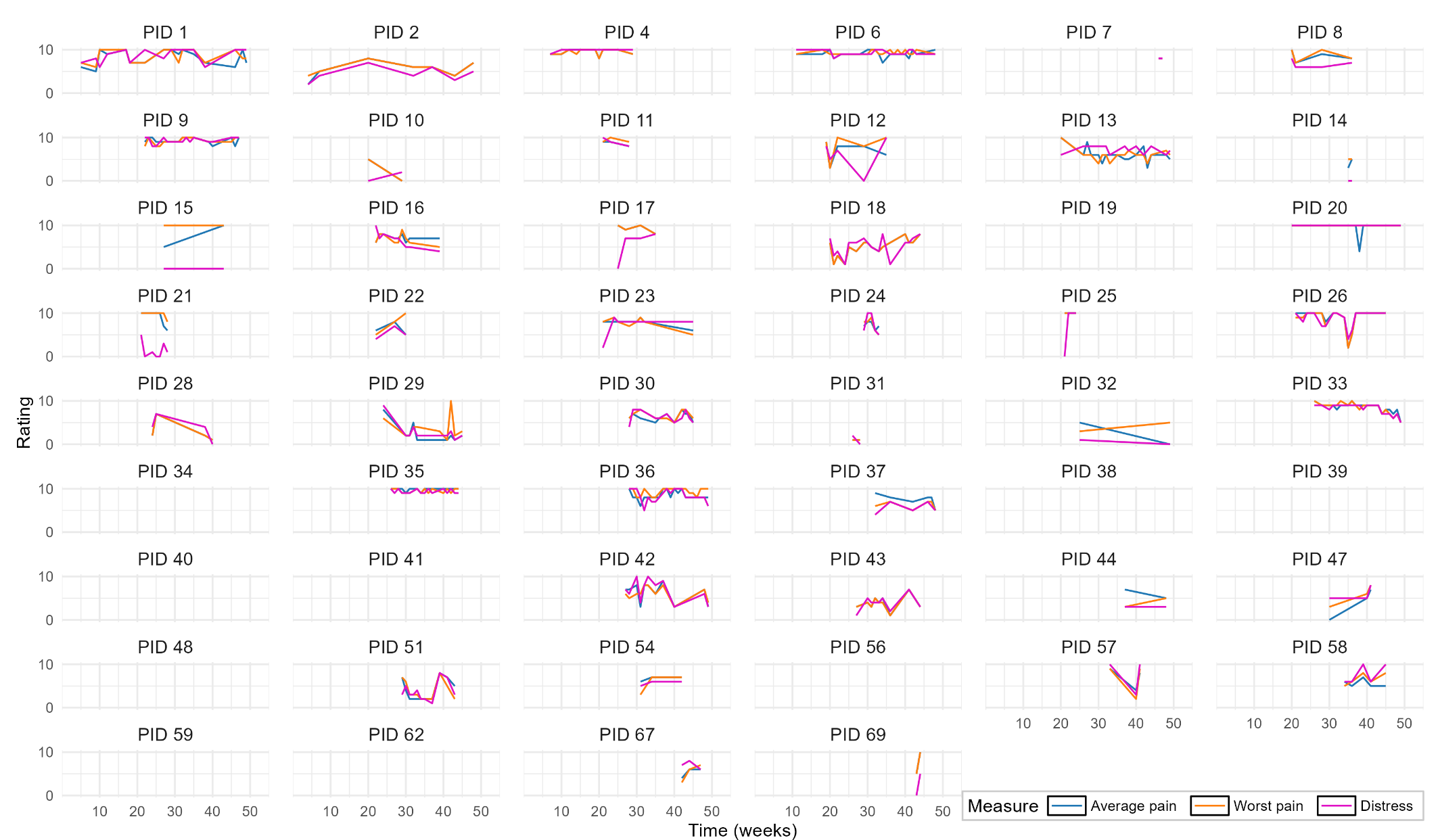

**Figure S10.** Pain and distress ratings over time for each participant id (PIDs), for all time points for which ratings had been provided for both distress and pain. Co-fluctuation was apparent for PIDs 42, 43, 44, 47, 51, 54, 57, 48. No lines in an individual plot mean distress or pain ratings were provided during a particular week but not in consecutive weeks. Blue lines show average pain, orange worst pain and magenta distress ratings over time.

#### Concurrent associations

##### Cumulative regression model diagnostics

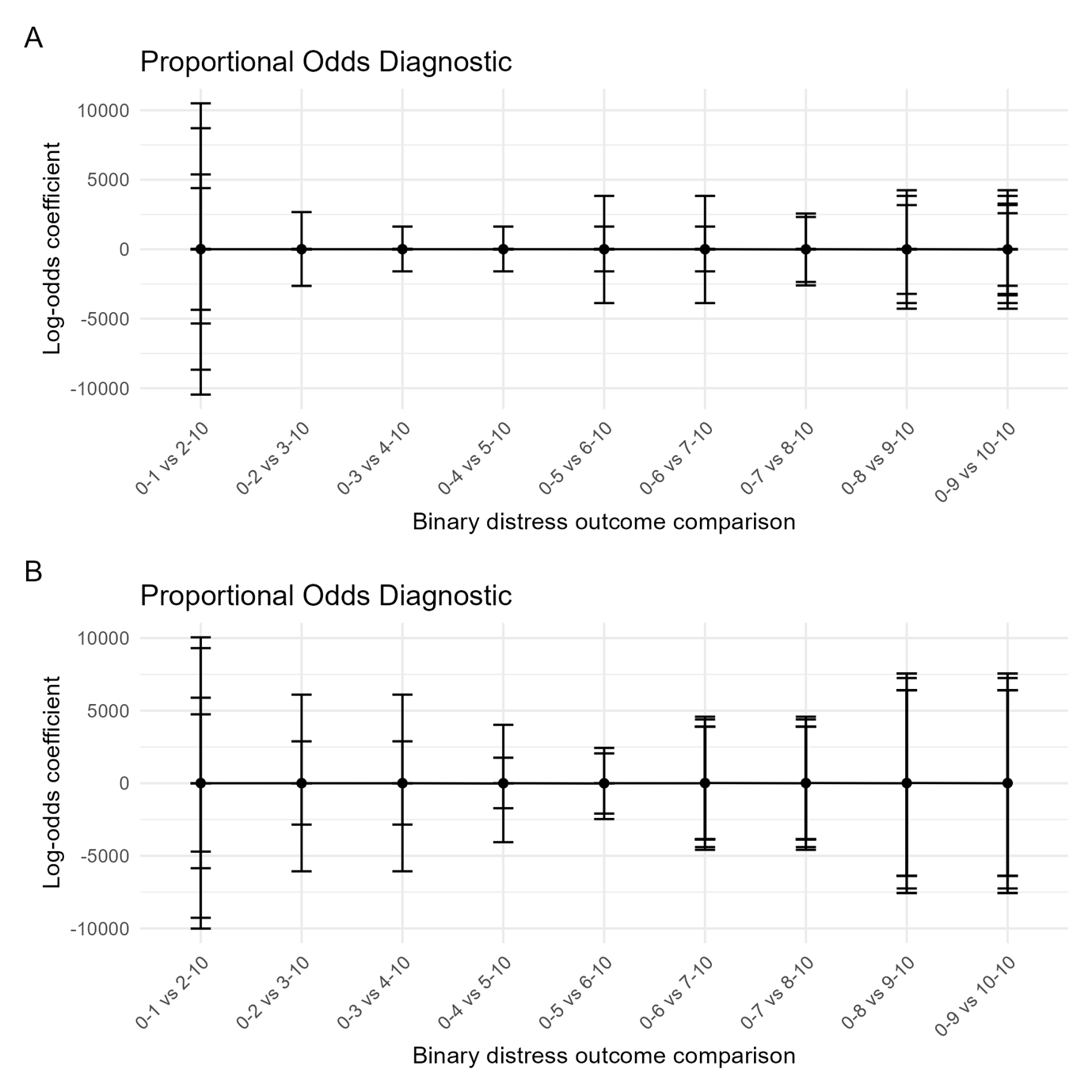

**Figure S11.** Proportional assumptions of the relationship between unstandardized, concurrently-reported distress (outcome) and a) worst pain intensity and b) average pain intensity (predictors). Dots indicate log odds estimates and lines 95% CIs.

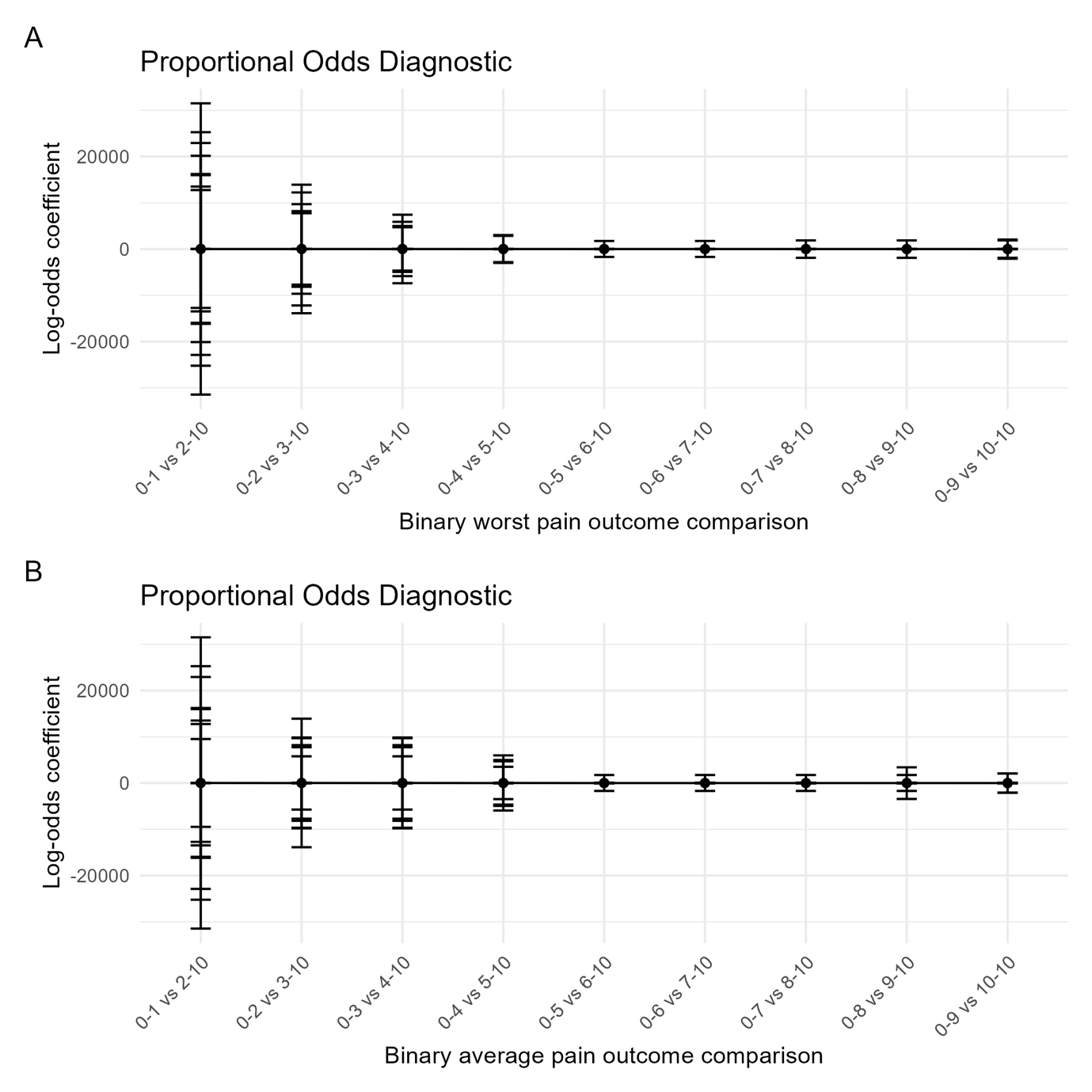

**Figure S12.** Proportional assumptions of the relationship between concurrently-reported distress (predictor) and unstandardized a) worst pain intensity and b) average pain intensity (outcomes). Dots indicate log odds estimates and lines 95% CIs.

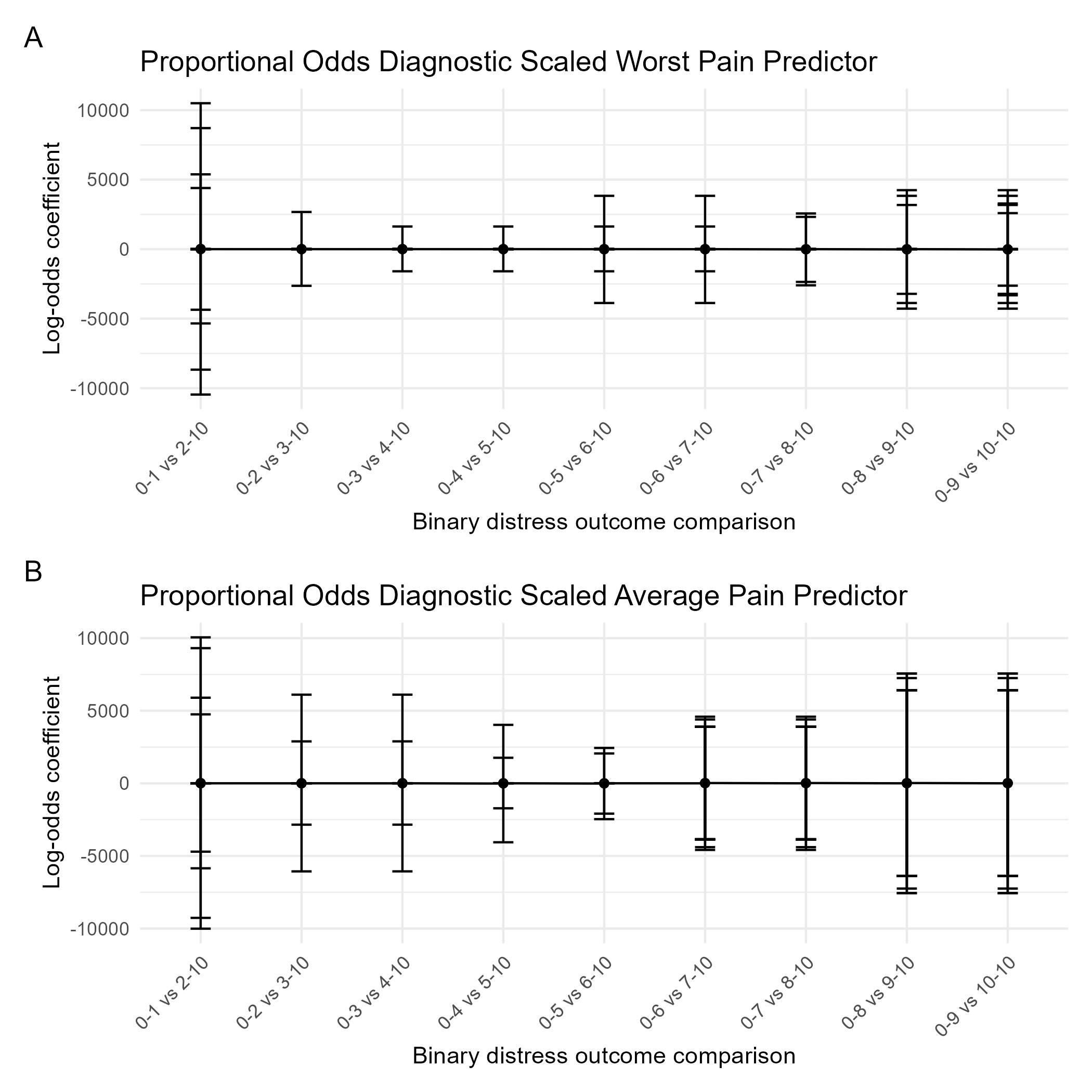

**Figure S13.** Proportional assumptions of the relationship between concurrently-reported distress (outcome) and standardized a) worst pain intensity and b) average pain intensity (predictor). Dots indicate log odds estimates and lines 95% CIs.

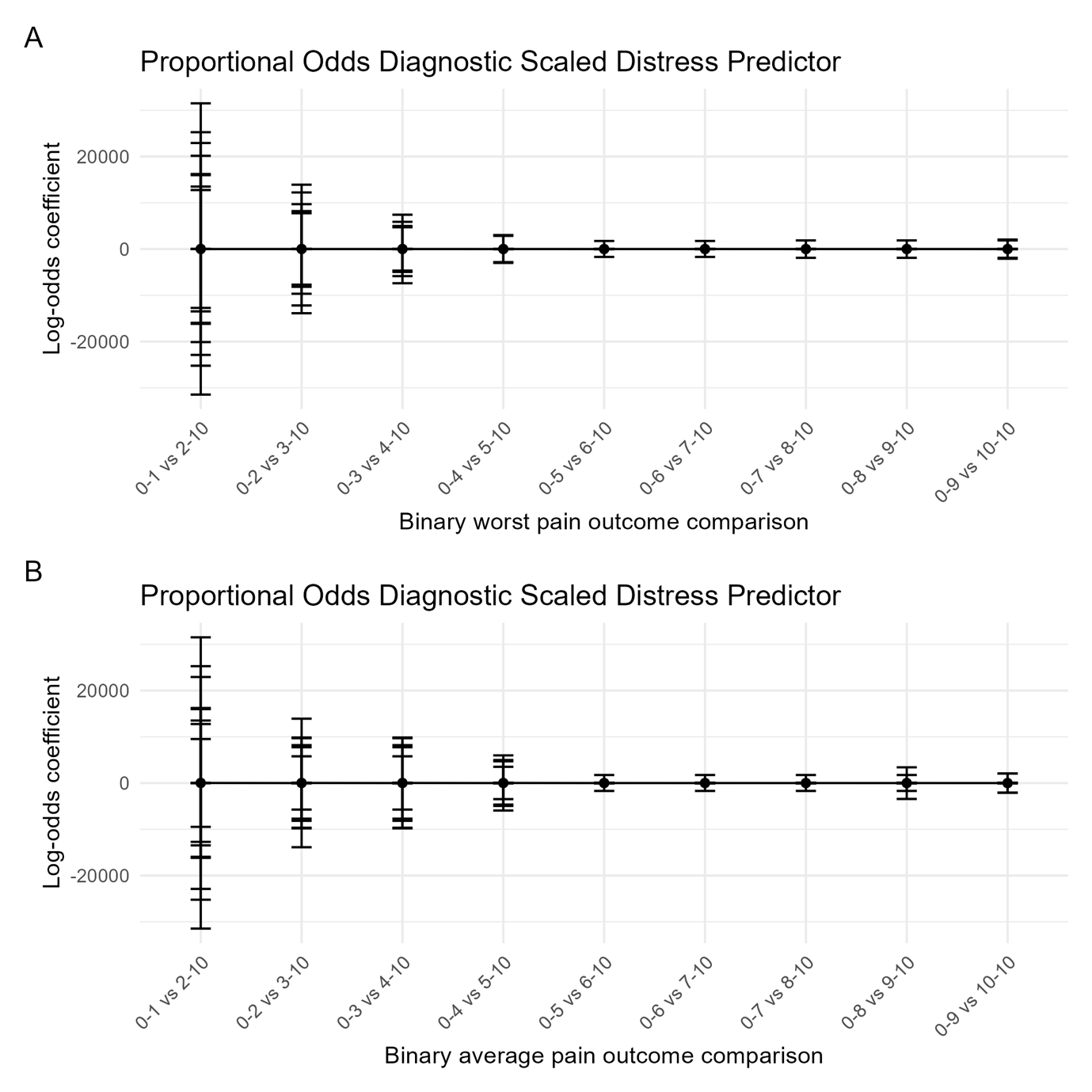

**Figure S14.** Proportional assumptions of the relationship between standardized distress (predictor) and a) worst pain intensity and b) average pain intensity (outcome). Dots indicate log odds estimates and lines 95% CIs.

##### Cumulative linear regression mixed models

| Table S3: Unadjusted ordinal regression models for concurrently reported distress (outcome) and pain intensity in the past week – unscaled variables | | | | | | |
| --- | --- | --- | --- | --- | --- | --- |
|  | **Worst pain as predictor** | | | **Average pain as predictor** | | |
| *Predictors* | *Odds Ratios* | *CI* | *p* | *Odds Ratios* | *CI* | *p* |
| worst pain rating | 2.08 | 1.77 – 2.43 | **<0.001** |  |  |  |
| average pain rating |  |  |  | 2.08 | 1.79 – 2.42 | **<0.001** |
| **Random Effects** | | | | | | |
| σ^2^ | 3.29 | | | 3.29 | | |
| τ_00_ | 9.66 _blinded_upi_id_ | | | 8.16 _blinded_upi_id_ | | |
| ICC | 0.75 | | | 0.71 | | |
| N | 52 _blinded_upi_id_ | | | 52 _blinded_upi_id_ | | |
| Observations | 368 | | | 367 | | |
| Marginal R^2^ / Conditional R^2^ | 0.220 / 0.802 | | | 0.241 / 0.782 | | |

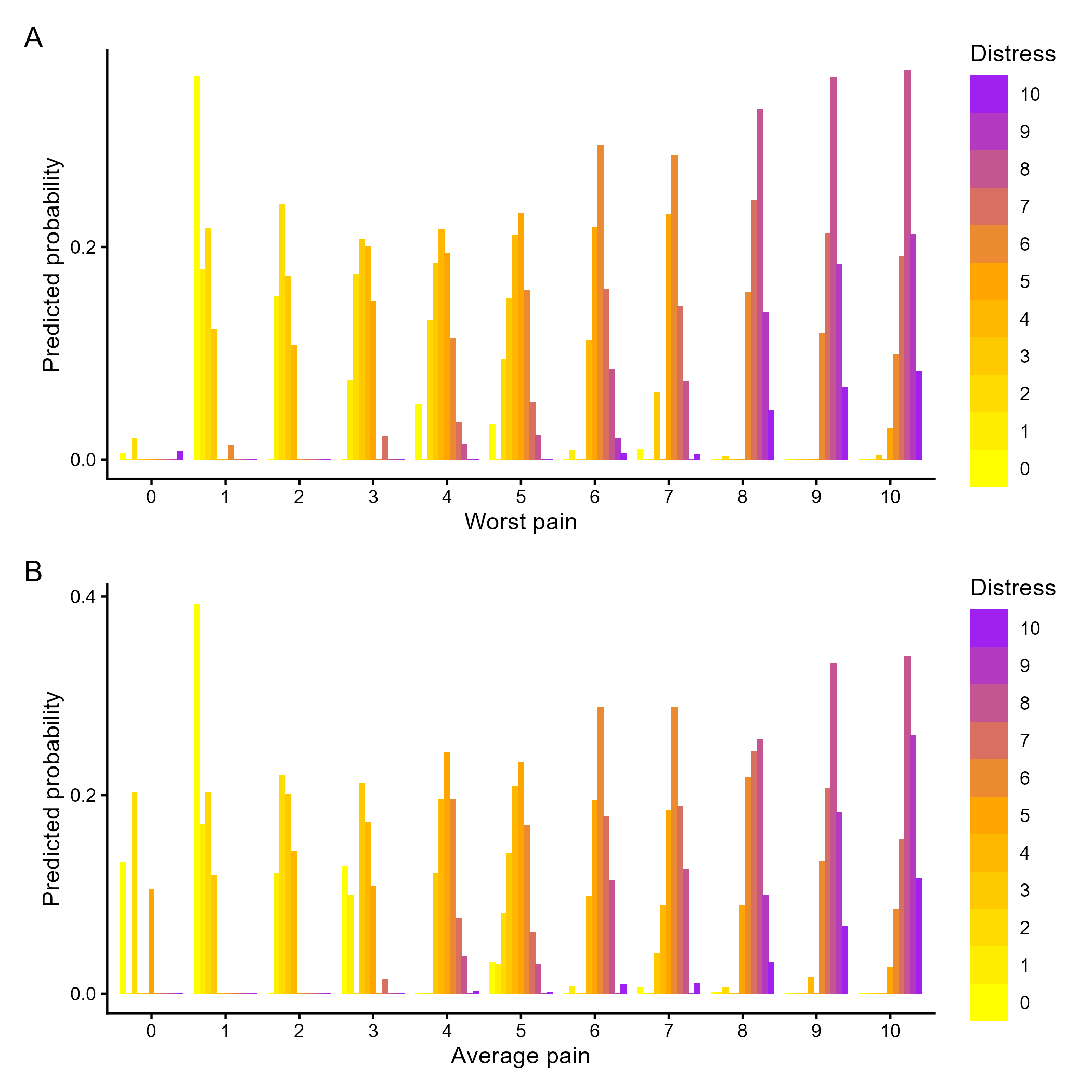

Figure S15: Statistical predictions of each level of distress (outcome) by each level of concurrent worst pain (A) or average pain (B). Ratings in the lower range of the pain scale most strongly predicted concurrent distress of 0, 2, or 3/10 (yellow bars); ratings in the middle range of the pain scale most strongly predicted distress of 6/10 (orange bars), and ratings in the upper range of the pain scale most strongly predicted distress of 8/10 (purple bars).

| Table S4: Unadjusted ordinal regression models for concurrently reported distress (outcome) and pain intensity in the past week - scaled predictor | | | | | | |
| --- | --- | --- | --- | --- | --- | --- |
|  | **Worst pain as predictor** | | | **Average pain as predictor** | | |
| *Predictors* | *Odds Ratios* | *CI* | *p* | *Odds Ratios* | *CI* | *p* |
| worst pain rating | 6.77 | 4.47 – 10.23 | **<0.001** |  |  |  |
| average pain rating |  |  |  | 6.77 | 4.57 – 10.03 | **<0.001** |
| **Random Effects** | | | | | | |
| σ^2^ | 3.29 | | | 3.29 | | |
| τ_00_ | 9.69 _blinded_upi_id_ | | | 8.22 _blinded_upi_id_ | | |
| ICC | 0.75 | | | 0.71 | | |
| N | 52 _blinded_upi_id_ | | | 52 _blinded_upi_id_ | | |
| Observations | 368 | | | 367 | | |
| Marginal R^2^ / Conditional R^2^ | 0.220 / 0.802 | | | 0.241 / 0.783 | | |

| Table S5: Unadjusted ordinal regression models for concurrently reported distress and pain intensity in the past week (outcome) – unscaled variables | | | | | | |
| --- | --- | --- | --- | --- | --- | --- |
|  | **Worst pain as outcome** | | | **Average pain as outcome** | | |
| *Predictors* | *Odds Ratios* | *CI* | *p* | *Odds Ratios* | *CI* | *p* |
| distress rating | 1.91 | 1.64 – 2.22 | **<0.001** | 1.85 | 1.61 – 2.13 | **<0.001** |
| **Random Effects** | | | | | | |
| σ^2^ | 3.29 | | | 3.29 | | |
| τ_00_ | 11.08 _blinded_upi_id_ | | | 5.52 _blinded_upi_id_ | | |
| ICC | 0.77 | | | 0.63 | | |
| N | 52 _blinded_upi_id_ | | | 52 _blinded_upi_id_ | | |
| Observations | 368 | | | 367 | | |
| Marginal R^2^ / Conditional R^2^ | 0.205 / 0.818 | | | 0.277 / 0.730 | | |

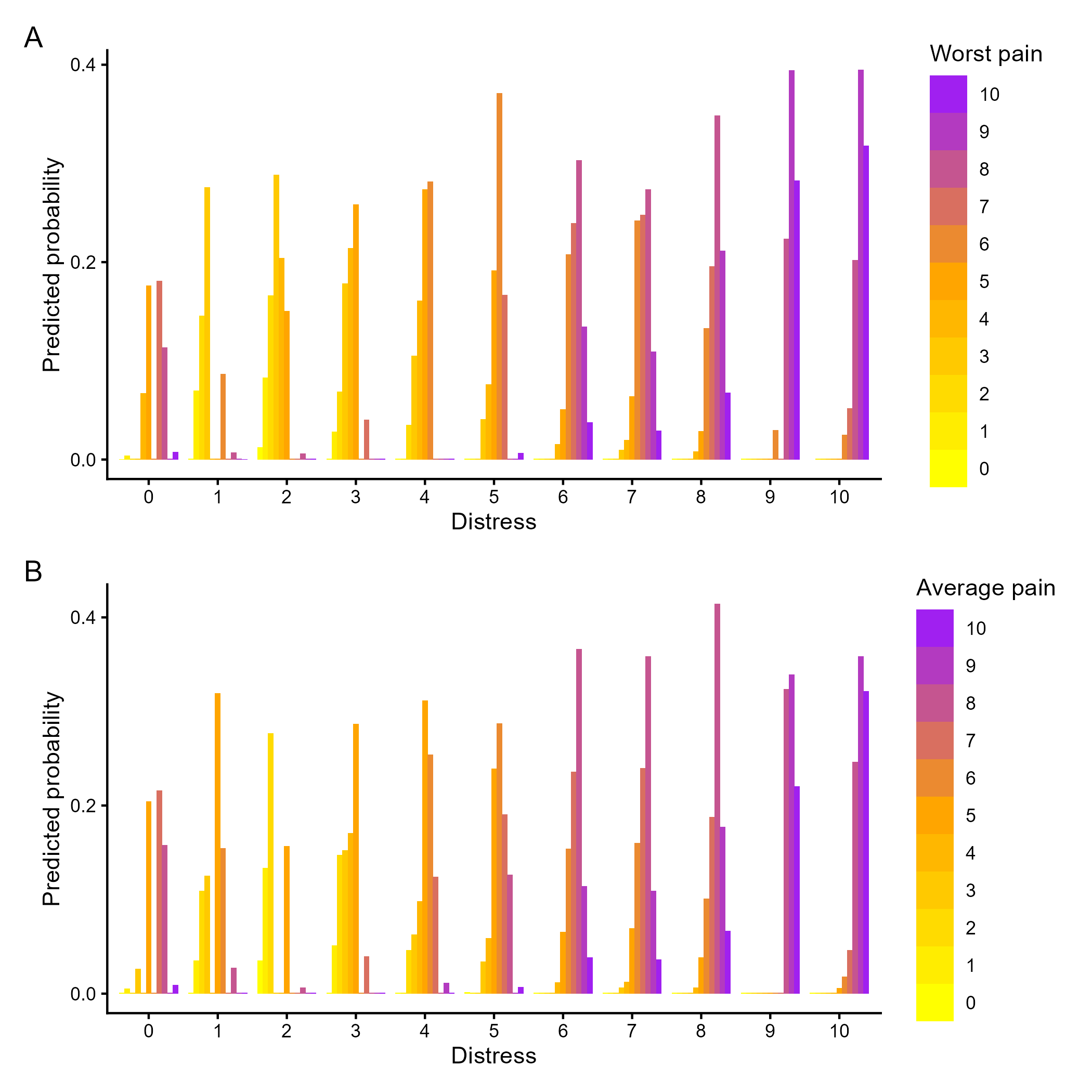

Figure S16: Statistical predictions of each level of concurrent worst pain (a) and average pain (b) ratings by each level of distress (predictor). Distress ratings of less than 3 most strongly predicted worst pain of 5, 6, 7, or 8/10; distress ratings of 3-5 most strongly predicted worst pain of 4, 5, or 6/10, and distress ratings of 6-10 most strongly predicted worst pain of 7, 8 or 9/10.

| Table S6: Unadjusted ordinal regression models for concurrently reported distress (predictor) and pain intensity in the past week (outcome) - scaled predictor | | | | | | |
| --- | --- | --- | --- | --- | --- | --- |
|  | **Worst pain as outcome** | | | **Average pain as outcome** | | |
| *Predictors* | *Odds Ratios* | *CI* | *p* | *Odds Ratios* | *CI* | *p* |
| distress rating | 11.57 | 6.48 – 20.63 | **<0.001** | 10.37 | 6.12 – 17.56 | **<0.001** |
| **Random Effects** | | | | | | |
| σ^2^ | 3.29 | | | 3.29 | | |
| τ_00_ | 11.08 _blinded_upi_id_ | | | 5.52 _blinded_upi_id_ | | |
| ICC | 0.77 | | | 0.63 | | |
| N | 52 _blinded_upi_id_ | | | 52 _blinded_upi_id_ | | |
| Observations | 368 | | | 367 | | |
| Marginal R^2^ / Conditional R^2^ | 0.205 / 0.818 | | | 0.277 / 0.730 | | |

#### Time-lagged associations

##### Cumulative regression model diagnostics

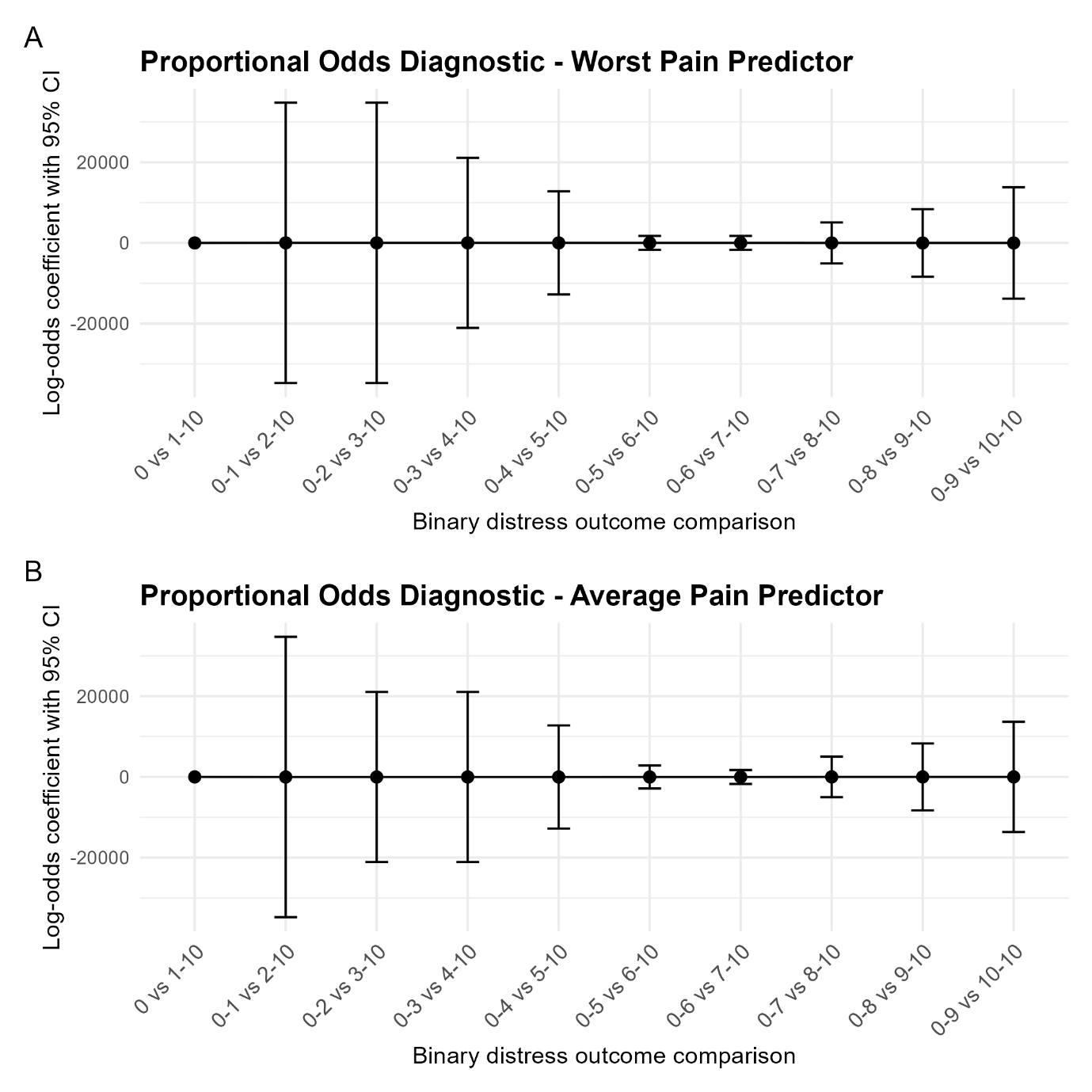
**Figure S17.** Proportional assumptions of the relationship between distress (outcome) and unstandardized a) worst pain intensity and b) average pain intensity (predictors). Dots indicate log odds estimates and lines 95% CIs.

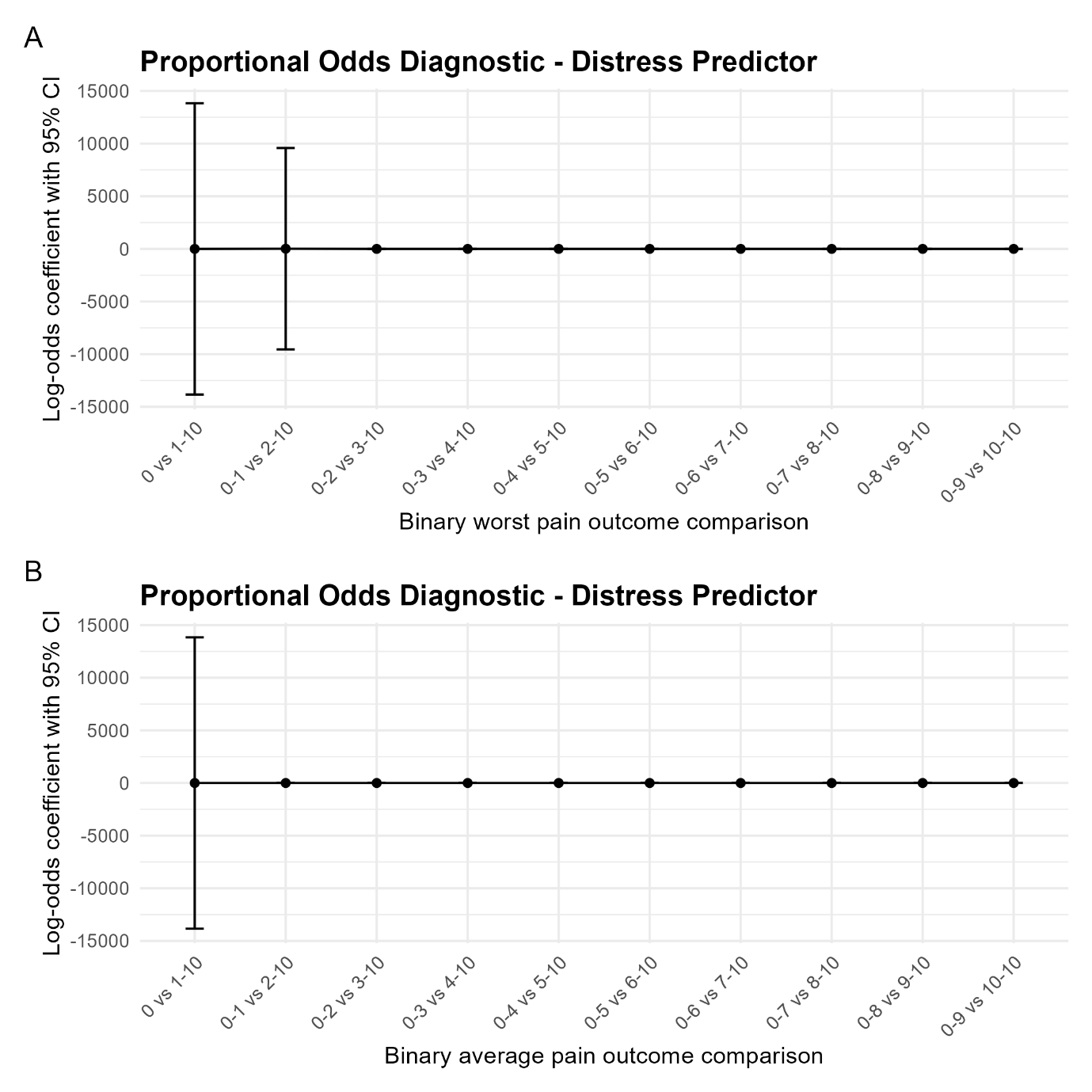

**Figure S18.** Proportional assumptions of the relationship between unstandardized distress (predictor) and a) worst pain intensity and b) average pain intensity (outcomes). Dots indicate log odds estimates and lines 95% CIs.

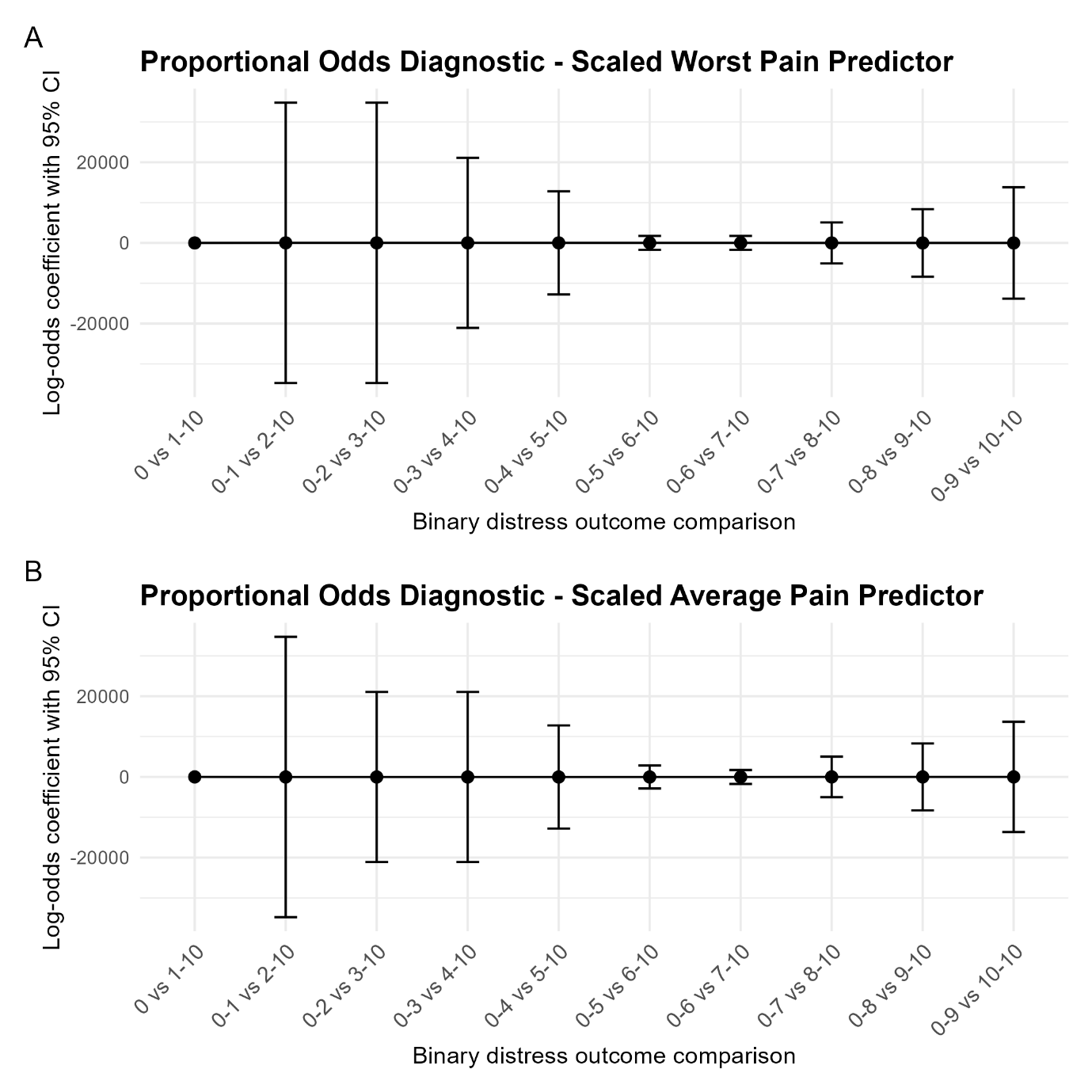
**Figure S19.** Proportional assumptions of the relationship between distress (outcome) and standardized a) worst pain intensity and b) average pain intensity (predictors). Dots indicate log odds estimates and lines 95% CIs.

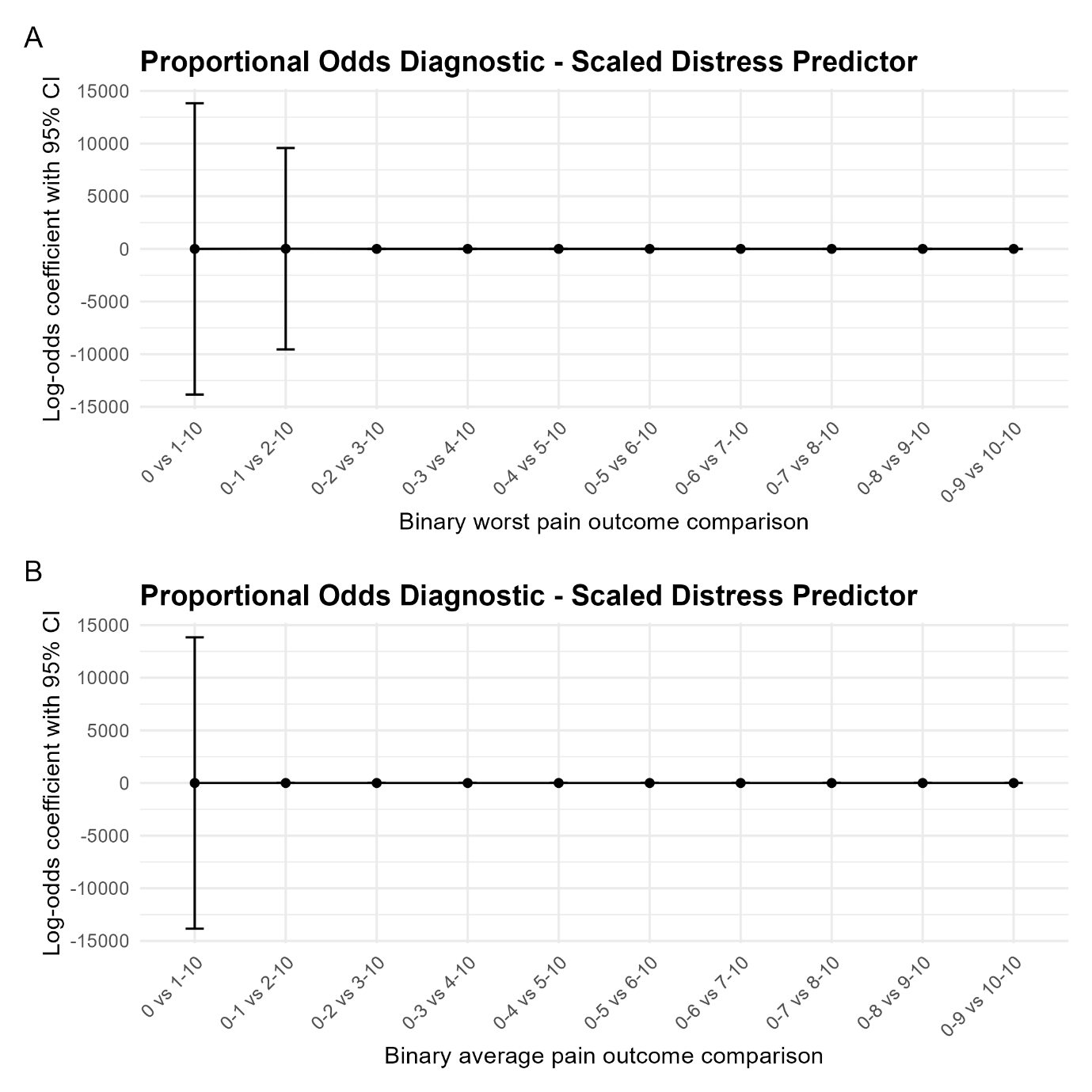
**Figure S20.** Proportional assumptions of the relationship between standardized distress (predictor) and a) worst pain intensity and b) average pain intensity (outcomes). Dots indicate log odds estimates and lines 95% CIs.

##### Cumulative linear regression mixed models

**Table S7.** Standardized unadjusted cumulative link mixed models for distress reported in the next week as dependent variable, and pain intensity ‘in the past week’, reported for the week before the distress report – i.e. does greater pain temporally precede greater distress?

|  | ^a^ **Worst pain as predictor** | | | ^a^ **Average pain as predictor** | | |
| --- | --- | --- | --- | --- | --- | --- |
| *Predictors* | *Odds Ratios* | *CI* | *p* | *Odds Ratios* | *CI* | *p* |
| Lag worst pain | 2.51 | 1.57 – 4.02 | **<0.001** |  |  |  |
| Lag average pain |  |  |  | 2.54 | 2.52 – 2.56 | **<0.001** |
| **Random Effects** | | | | | | |
| σ^2^ | 3.29 | | | 3.29 | | |
| τ_00_ | 12.78 _blinded_upi_id_ | | | 13.58 _blinded_upi_id_ | | |
| ICC | 0.80 | | | 0.81 | | |
| N | 45 _blinded_upi_id_ | | | 45 _blinded_upi_id_ | | |
| Observations | 245 | | | 244 | | |
| Marginal R^2^ / Conditional R^2^ | 0.050 / 0.806 | | | 0.049 / 0.815 | | |

^a^ Worst pain and average pain predicted distress in the next week in the standardized CLMMs. On average, a 1-point increase in worst pain or average pain was associated with a 2.51 (95% CI: 1.57-4.02, p<0.001) or 2.54 (95% CI: 2.52-2.56, p<0.001) times the odds of a higher distress score the next week, respectively.

**Table S8.** Standardized unadjusted cumulative link mixed models with distress reported in the preceding week as the independent variable, and worst or average pain intensity ‘in the past week’ as dependent variable – i.e. does greater distress temporally precede greater pain the next week?

|  | ^a^ **Worst pain as outcome** | | | ^a^ **Average pain as outcome** | | |
| --- | --- | --- | --- | --- | --- | --- |
| *Predictors* | *Odds Ratios* | *CI* | *p* | *Odds Ratios* | *CI* | *p* |
| Lag distress | 2.36 | 1.28 – 4.34 | **0.017** | 1.94 | 1.04 – 3.63 | 0.114 |
| **Random Effects** | | | | | | |
| σ^2^ | 3.29 | | | 3.29 | | |
| τ_00_ | 7.61 _blinded_upi_id_ | | | 5.88 _blinded_upi_id_ | | |
| ICC | 0.70 | | | 0.64 | | |
| N | 47 _blinded_upi_id_ | | | 47 _blinded_upi_id_ | | |
| Observations | 255 | | | 255 | | |
| Marginal R^2^ / Conditional R^2^ | 0.043 / 0.711 | | | 0.031 / 0.653 | | |

^a^ Distress predicted worst pain, but not average pain in the standardized CLMMs. On average, a 1-point increase in distress was statistically significantly associated with 2.36 (95% CI: 1.28–4.34, p=0.017) the odds that worst pain intensity would be at a higher level in the next week.

#### Sensitivity analyses

##### Concurrent associations

| Table S9: Unadjusted ordinal regression models for concurrently reported distress (outcome) and pain intensity in the past week - sensitivity analysis without 22 responses with discrepant participant identifier | | | | | | |
| --- | --- | --- | --- | --- | --- | --- |
|  | **Worst pain predictor** | | | **Average pain predictor** | | |
| *Predictors* | *Odds Ratios* | *CI* | *p* | *Odds Ratios* | *CI* | *p* |
| Worst pain rating | 2.11 | 2.10 – 2.13 | **<0.001** |  |  |  |
| Average pain rating |  |  |  | 2.12 | 1.81 – 2.49 | **<0.001** |
| **Random Effects** | | | | | | |
| σ^2^ | 3.29 | | | 3.29 | | |
| τ_00_ | 9.15 _blinded_upi_id_ | | | 9.15 _blinded_upi_id_ | | |
| ICC | 0.76 | | | 0.74 | | |
| N | 52 _blinded_upi_id_ | | | 52 _blinded_upi_id_ | | |
| Observations | 358 | | | 357 | | |
| Marginal R^2^ / Conditional R^2^ | 0.219 / 0.814 | | | 0.239 / 0.799 | | |

| Table S10: Unadjusted concurrent ordinal regression models for concurrently reported distress and pain intensity in the past week (outcome) - sensitivity analysis without 22 responses with discrepant participant identifier | | | | | | |
| --- | --- | --- | --- | --- | --- | --- |
|  | **Worst pain outcome** | | | **Average pain outcome** | | |
| *Predictors* | *Odds Ratios* | *CI* | *p* | *Odds Ratios* | *CI* | *p* |
| distress rating | 1.98 | 1.68 – 2.33 | **<0.001** | 1.89 | 1.63 – 2.19 | **<0.001** |
| **Random Effects** | | | | | | |
| σ^2^ | 3.29 | | | 3.29 | | |
| τ_00_ | 12.19 _blinded_upi_id_ | | | 6.90 _blinded_upi_id_ | | |
| ICC | 0.79 | | | 0.68 | | |
| N | 52 _blinded_upi_id_ | | | 52 _blinded_upi_id_ | | |
| Observations | 358 | | | 357 | | |
| Marginal R^2^ / Conditional R^2^ | 0.210 / 0.832 | | | 0.258 / 0.761 | | |

##### Time-lagged associations

| Table S11: Unadjusted ordinal regression models for distress (outcome) and pain intensity ‘in the past week’, reported the week before the distress rating) – i.e. does greater pain temporally precede greater distress? Sensitivity analysis without 22 responses with discrepant participant identifiesr. | | | | | | |
| --- | --- | --- | --- | --- | --- | --- |
|  | **Worst pain predictor** | | | **Average pain predictor** | | |
| *Predictors* | *Odds Ratios* | *CI* | *p* | *Odds Ratios* | *CI* | *p* |
| Lag worst pain | 1.40 | 1.17 – 1.68 | **<0.001** |  |  |  |
| Lag average pain |  |  |  | 1.44 | 1.19 – 1.73 | **0.001** |
| **Random Effects** | | | | | | |
| σ^2^ | 3.29 | | | 3.29 | | |
| τ_00_ | 13.70 _blinded_upi_id_ | | | 14.66 _blinded_upi_id_ | | |
| ICC | 0.81 | | | 0.82 | | |
| N | 44 _blinded_upi_id_ | | | 44 _blinded_upi_id_ | | |
| Observations | 238 | | | 237 | | |
| Marginal R^2^ / Conditional R^2^ | 0.045 / 0.815 | | | 0.048 / 0.826 | | |

| Table S12: Unadjusted ordinal regression models for distress reported in the preceding week, and pain intensity ‘in the past week’ (outcome) – i.e. does greater distress temporally precede greater pain? Sensitivity analysis without 22 responses with discrepant participant identifiers. | | | | | | |
| --- | --- | --- | --- | --- | --- | --- |
|  | **Worst pain outcome** | | | **Average pain outcome** | | |
| *Predictors* | *Odds Ratios* | *CI* | *p* | *Odds Ratios* | *CI* | *p* |
| Lag distress | 1.26 | 1.07 – 1.49 | **0.035** | 1.16 | 1.15 – 1.17 | **<0.001** |
| **Random Effects** | | | | | | |
| σ^2^ | 3.29 | | | 3.29 | | |
| τ_00_ | 7.34 _blinded_upi_id_ | | | 6.76 _blinded_upi_id_ | | |
| ICC | 0.69 | | | 0.67 | | |
| N | 46 _blinded_upi_id_ | | | 46 _blinded_upi_id_ | | |
| Observations | 246 | | | 246 | | |
| Marginal R^2^ / Conditional R^2^ | 0.047 / 0.705 | | | 0.020 / 0.679 | | |
